# Bridging surveillance gaps in dengue: a hierarchical model integrating mixed data sources for transmission estimation and vaccine targeting

**DOI:** 10.64898/2026.07.15.26358208

**Authors:** Bimandra A Djaafara, Iqbal RF Elyazar, Prima Yosephine, Asik Surya, Fadjar SM Silalahi, Agus Handito, Burhannudin Thohir, Desfalina Aryani, Dyana Gunawan, Anzala Khoirun Nisa, Edi Prianto, Iriani Samad, Alex R Cook, Angkana T Huang, Hannah E Clapham, Samir Bhatt, Swapnil Mishra

## Abstract

Estimating dengue force of infection (FOI) is essential for understanding transmission dynamics and targeting intervention programmes, yet the surveillance data in endemic settings required for estimation are often incomplete, with varying formats. We developed a Bayesian hierarchical catalytic model that jointly fits age-stratified case data, aggregate case data, and seroprevalence surveys within a single framework, incorporating external covariates to improve parameter identifiability. Synthetic validation showed that FOI and seroprevalence were recovered with little bias even when most districts contributed only aggregate case data, whereas reporting rates were reliably recovered only where age-stratified case data were available. Applied to 128 districts across Java and Bali, Indonesia (2016–2024), the model revealed substantial spatial heterogeneity in FOI and reporting rates. Many districts in Java exceeded the WHO-suggested seroprevalence threshold for vaccine introduction, yet were classified as low-priority when using reported incidence as prioritisation criterion, particularly in areas with weak surveillance. Model-based seroprevalence estimation, integrating multiple data sources, offers a more consistent basis for identifying high-priority districts for vaccine introduction, and is less susceptible to surveillance bias than reported incidence alone.

## INTRODUCTION

Dengue is among the most rapidly expanding mosquito-borne viral diseases, with cases reported globally to the World Health Organization (WHO) increasing from ∼500 thousand in 2000 to around ∼14.6 million in 2024 across over 100 countries (*1*). However, reported case counts substantially underestimate the true burden of dengue (*2*, *3*), as the majority of infections go undetected by routine surveillance systems. Interpretation is complicated by reporting rates that vary widely across settings (*3*, *4*). Estimating the force of infection (FOI), the rate at which susceptible individuals become infected, is critical to understanding transmission dynamics and guiding intervention strategies. FOI estimates underpin key policy decisions, including the targeting of dengue vaccination programmes of recently licensed dengue vaccines, which the WHO recommends for use in high transmission intensity settings (*5*). Seroprevalence at age 9 has been suggested as one indicator of transmission intensity to guide these decisions, with a threshold of 60% proposed to identify high-transmission areas (*5*).

In practice, FOI estimation is constrained by the quality and completeness of available surveillance data. Many endemic countries, particularly in Southeast Asia, rely on routine surveillance systems that capture only a fraction of infections, with reporting rates that vary substantially across locations (*3*) and contain several sources of bias. In Indonesia, the world’s fourth most populous country, age-stratified case data, which provide the richest information for FOI estimation, are available in only a subset of districts or provinces, while others report only aggregate annual totals. Seroprevalence surveys offer complementary information on cumulative exposure but are costly and resource-intensive (*6*), limiting their spatial and temporal coverage. This heterogeneity in data availability presents a fundamental challenge: how to estimate FOI consistently across locations when the underlying data differ in format, completeness, and quality.

Catalytic compartment models provide an established framework for estimating FOI from age-structured epidemiological data (*7–12*) and seroprevalence survey data (*12–14*). However, existing approaches are typically constructed around the assumption of uniform age-stratified data across all locations and estimate FOI for individual locations independently, limiting their applicability in settings with mixed data types. Recent work has extended catalytic models to accommodate multiple locations within hierarchical structures (*10*), but the joint integration of heterogeneously structured data—case data that are age-stratified in some locations, but aggregate in others, seroprevalence estimates with different age categorisations across studies, and external covariates with different spatial resolutions—within a single inference framework that appropriately pools information across all locations has not been attempted. Such models are important for rigorous data synthesis and addressing policy-relevant questions.

Here, we present a Bayesian hierarchical catalytic model that addresses these limitations. The model estimates time-varying FOI across 128 districts in seven provinces of Java and Bali, Indonesia, across 2016–2024, jointly fitting age-stratified and aggregate case data alongside seroprevalence surveys within a two-level hierarchical structure. External covariates are incorporated to inform heterogeneity in both FOI and reporting rates, providing additional structure to aid parameter identifiability. We validated the framework using synthetic data under four scenarios of decreasing data completeness, with covariates explaining only a small share (10%) of between-district variation in FOI, as estimated for the real data, and assessed sensitivity for higher shares. To evaluate the contribution of each data source, we fit four model variants—a full model with covariates and seroprevalence, and three reduced models omitting covariates, seroprevalence, or both—to both synthetic and real data. We characterised spatial and temporal patterns of dengue transmission in Java and Bali, and used the resulting seroprevalence estimates to assess district-level vaccine eligibility under WHO-suggested criteria.

## RESULTS

### Synthetic data validation

The model recovered true parameter values well across all four data completeness scenarios in a setting where covariates explained only a small share (10%) of between-district variation in FOI, comparable to that estimated for Java and Bali data (**Fig. 1**; see **Materials and Methods**). Each scenario was fitted with and without covariates, where covariates with known ‘true’ effects on FOI and reporting rates were specified in the data-generating processes. The scenarios span progressively coarser age stratification: all five simulated provinces reporting nine age groups (S1); all reporting four age groups (S2); two provinces reporting nine age groups and three reporting four age groups (S3); and a mixed setting in which three of five provinces report aggregate annual totals only (S4), which most closely mirrors the Indonesian data (**Table 1**). FOI estimates closely tracked true values across provinces and years, with similar accuracy in models with and without covariates (**Fig. 1A**). Reporting rate estimates were well recovered in S1-S3 but were systematically underestimated in S4, where most districts report only aggregate case data, with estimates plateauing at around 18% for districts whose true reporting rates exceeded 20% (**Fig. 1B**).

**Figure 1.**
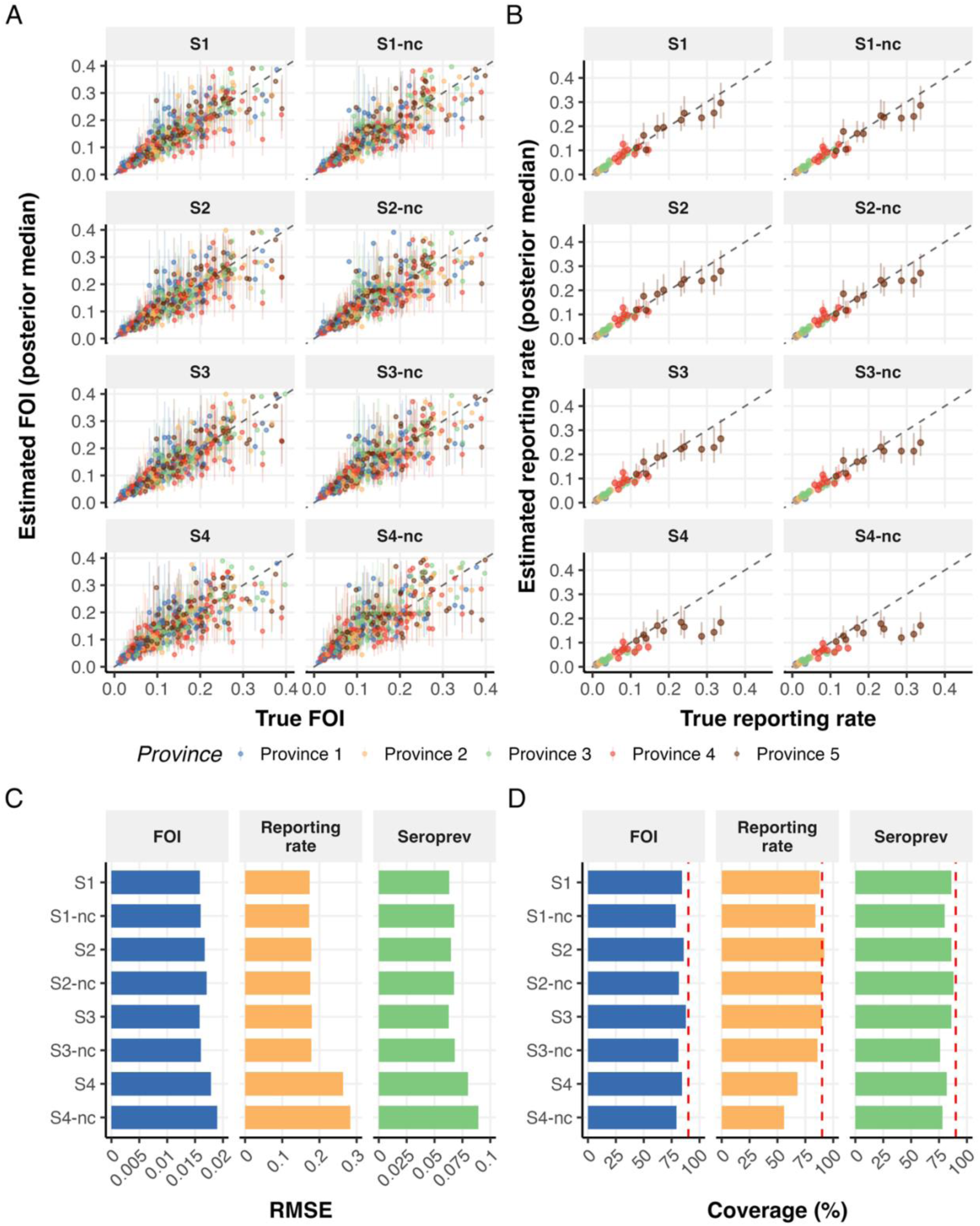
Synthetic data validation: parameter recovery across four data completeness scenarios (S1–S4), each fitted with and without covariates (-nc), in a setting where covariates explain ∼10% of between-district variation in FOI. **(A)** True versus estimated total FOI (summed over the four serotypes) across districts and years (posterior median with 90% CrI), coloured by provinces. **(B)** True versus estimated reporting rate across districts. **(C)** Root mean squared error (RMSE) for FOI, reporting rate (relative RMSE), and seroprevalence at age 9 at the end of 2024. **(D)** Coverage of 90% credible intervals; dashed lines indicate the nominal 90% coverage. S1: all districts with 9 age groups; S2: all with 4 age groups; S3: mixed 9 and 4 age groups; S4: mixed age-stratified and aggregate-only districts. Dashed diagonal lines in A-B indicate perfect agreement. Note that x-axis scales differ across parameter types in panel C, reflecting differences in parameter ranges.

**Table 1.**
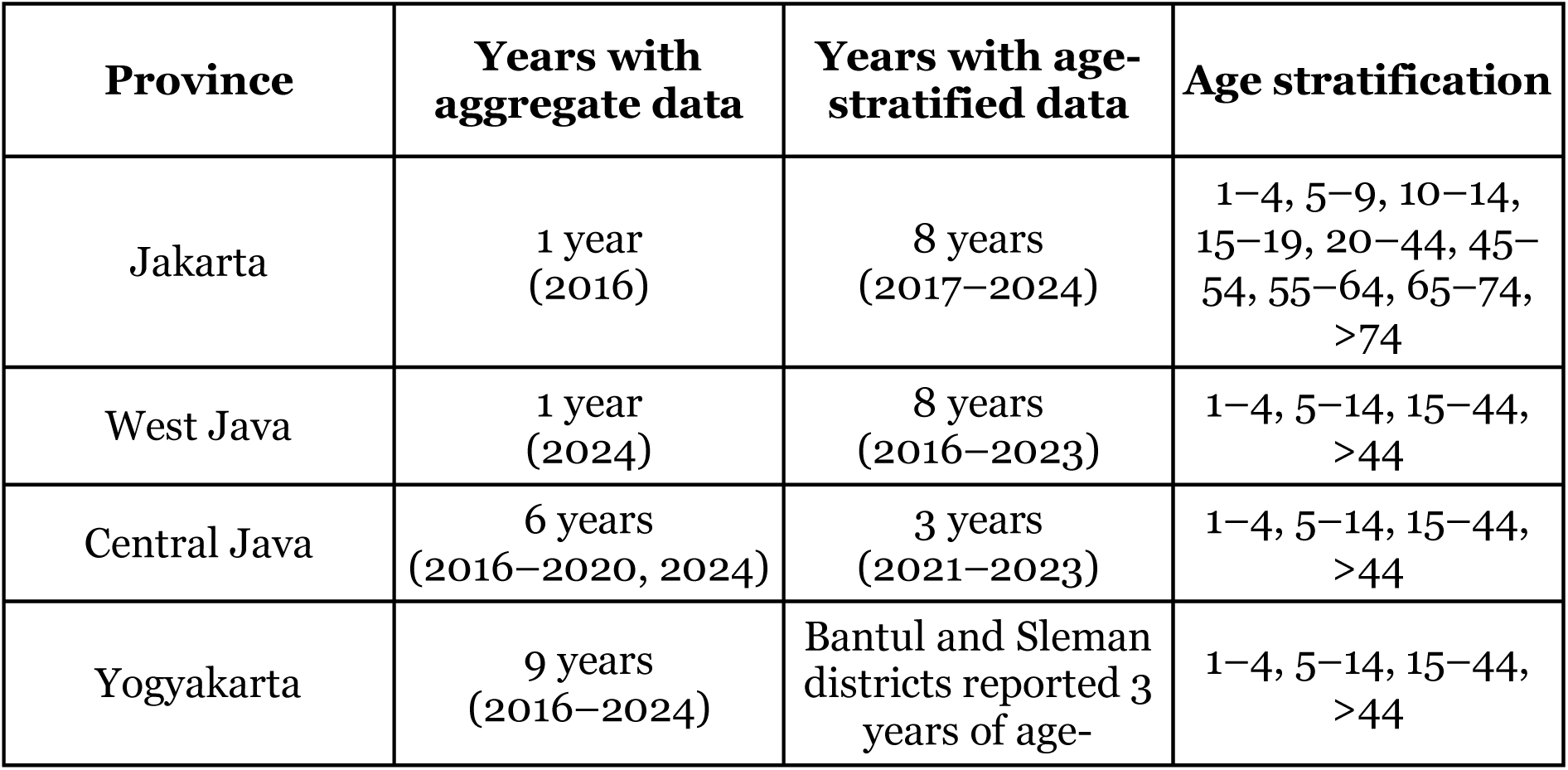
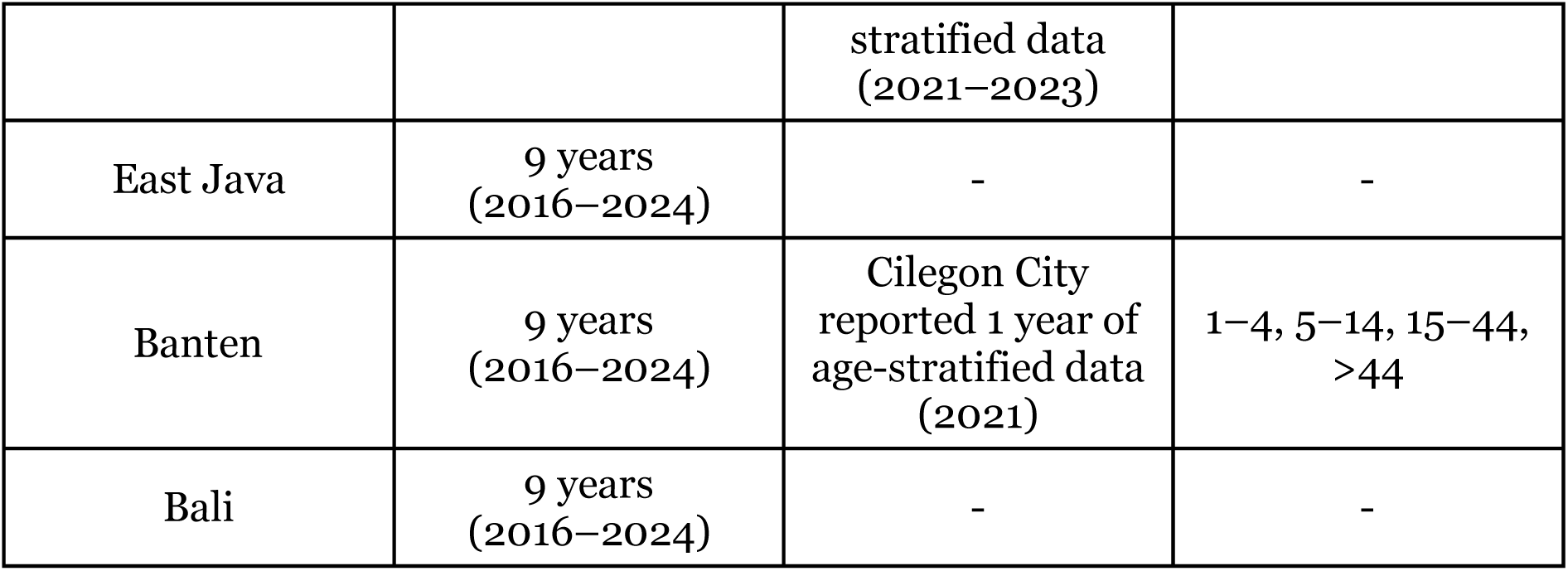
Dengue surveillance data availability and format by province, 2016-2024.

Across scenarios, the completeness of age-stratified data mattered most for separating reporting rates from FOI (**Fig. 1C**). The error in reporting rate estimates was similar in S1-S3 (relative root mean square error, RMSE, 0.17-0.18), indicating that coarser age grouping cost little, but increased sharply in S4 (0.26). In contrast, FOI and seroprevalence at age 9 remained reasonably well estimated in S4, with RMSE only moderately higher than in S1-S3 (FOI: 0.018 versus 0.016-0.017; seroprevalence: 0.080 versus 0.063-0.065). Covariates had little effect on the accuracy of FOI point estimates, but their inclusion made the credible intervals more reliable (coverage of 90% CrI 84-88% with covariates, compared with 79-82% without, **Fig. 1D**). A similar gain was seen for reporting rates, and, in most scenarios, seroprevalence. Reporting rate coverage in S4 remained well below other scenarios with covariates (68%, and 56% without), consistent with the shrinkage of high reporting rates described above.

Accuracy of seroprevalence estimates at age 9 depended on how much a cohort’s exposure fell within the surveillance period (**Fig. 2A**). In S1-S3, RMSE was highest in 2016, when eight of the nine years of exposure precede the case data availability and rely on reconstructed historical FOI. Then it fell to its lowest around 2019 (S1: 0.065 to 0.034) and rose again towards 2024 (0.063), as FOI in the final years is informed only by cases in those years, without the additional constraint that later case data provide for earlier years. In S4, RMSE remained higher and nearly constant throughout (0.07-0.08). Model without covariates had slightly higher error in all scenarios, particularly in later years. Predicted seroprevalence at ages 5, 9 and 15 in 2024 closely matched true values in model with covariates, with little systematic bias and credible intervals that captured most true values (**Fig. 2B**). Without covariates, estimates tended to be biased upward, most notably in S4. Covariate coefficients were generally well recovered, although some FOI coefficients were biased in a consistent direction across all scenarios (**Fig. 2C**). These biases largely disappeared when covariates explained a larger share of between-district variation (**Fig. S1**), indicating that individual coefficients are weakly identified when covariate-driven heterogeneity is low, even though FOI itself is well estimated.

**Figure 2.**
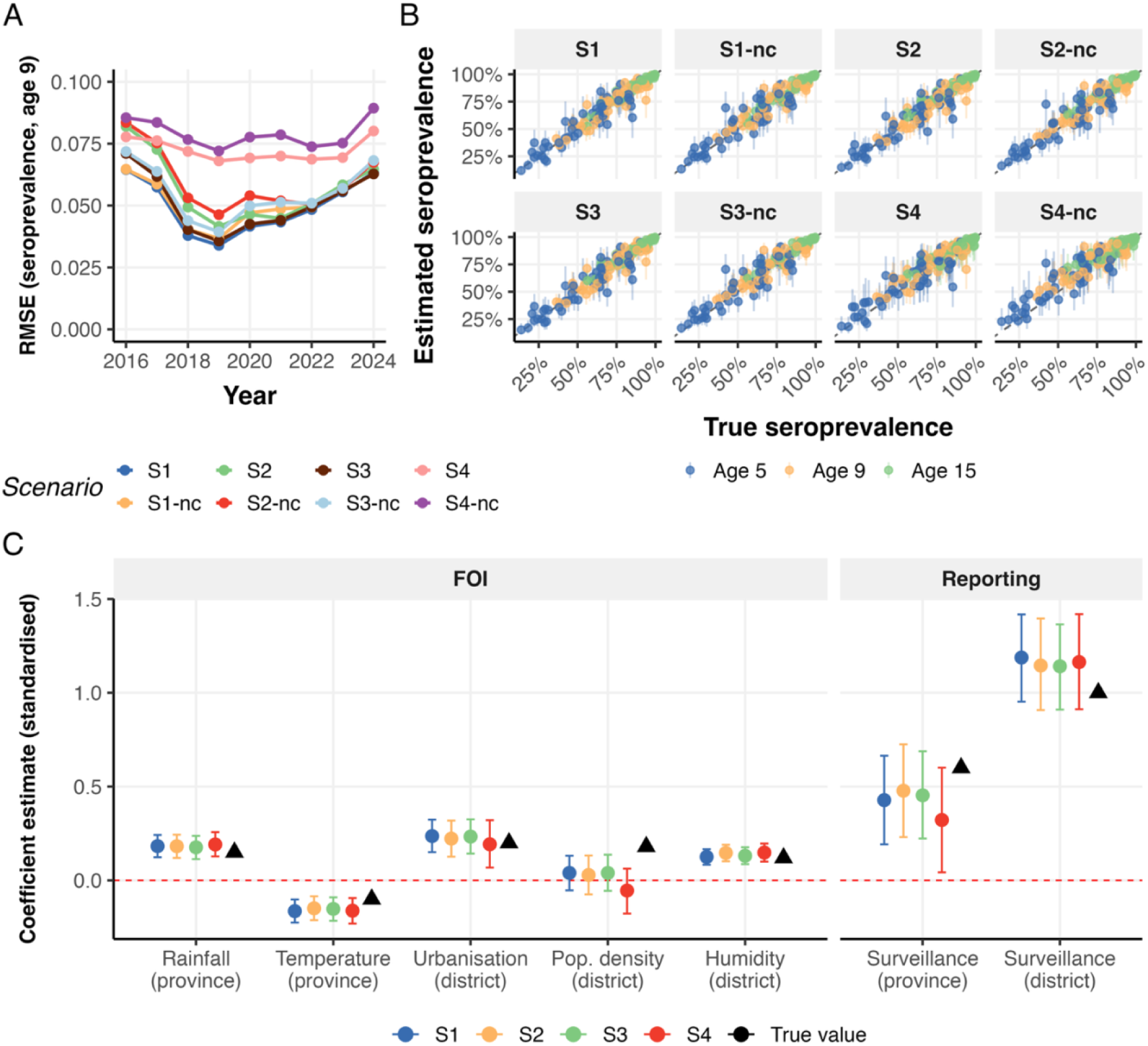
Seroprevalence accuracy over time and covariate recovery in synthetic validation. **(A)** RMSE of estimated seroprevalence at age 9 at the end of each year, 2016-2024. **(B)** Estimated versus true seroprevalence at ages 5, 9, and 15 at the end of 2024 (posterior median with 90% CrI); dashed lines indicate perfect agreement. **(C)** Standardised covariate coefficient recovery for FOI (left) and reporting rate (right), in the scenario with covariates; points and bars show posterior median and 90% CrI, and triangles indicate true values.

We conducted several sensitivity analyses to assess the robustness of these results. As covariates explained a larger share of between-district variation in FOI, accuracy and coverage improved substantially, while reporting rates in S4 remained the least accurate at every level (**Fig. S1**). Estimate bias was consistently the highest in S4, where shrinkage in reporting rates towards the shared mean led to their underestimation and, on average, led to overestimation of FOI and seroprevalence (**Fig. S2**). Adding a single seroprevalence survey, at the end of either 2014 or 2019, mainly improved seroprevalence estimates for years whose exposure depended on historical FOI, with little effect on estimates for 2024 (**Fig. S3-S7**). The model was also robust to covariate misspecification, as an uninformative covariate was correctly estimated as having no effect (**Fig. S8**) and including only a subset of the true covariates yielded performance between that of the full- and no-covariate models (**Fig. S9-S11**).

Overall, the synthetic validation demonstrates that, even when covariates explain only a small share of between-district variation, the hierarchical catalytic model recovers FOI and seroprevalence with little bias and high CrI coverage across data completeness scenario, while reporting rates are reliably recovered only where age-stratified case data are available.

### Application to Java and Bali data, 2016-2024

The study area comprised 128 districts across seven provinces in Java—the world’s most populous island—and Bali (**Fig. 3A**). West Java reported the highest absolute case counts, while Bali had the highest reported incidence rate per 100,000 population (**Fig. 3B-C**). Despite differences in magnitude, all provinces showed broadly synchronous outbreak timing, with peaks in 2016, 2019, 2022, and 2024. Seroprevalence data were available from surveys conducted in 2014, 2016, 2019, 2020, and 2024 within the study area (**Fig. 3D**). Only the 2014 nationwide urban serosurvey, covering 22 sites in Java and Bali, was included for model fitting, where it informs historical FOI through the reconstruction of historical immunity. The remaining surveys, from Kota Yogyakarta (2016, 2019, and 2024) and Kota Denpasar (2020), were held out for out-of-sample validation. Full model specification, prior choices, and covariate selection rationale are detailed in **Materials and Methods**. Unless otherwise stated, results are from the full model (covariates and seroprevalence included). Model comparison across all four variants is presented at the end of this section.

**Figure 3.**
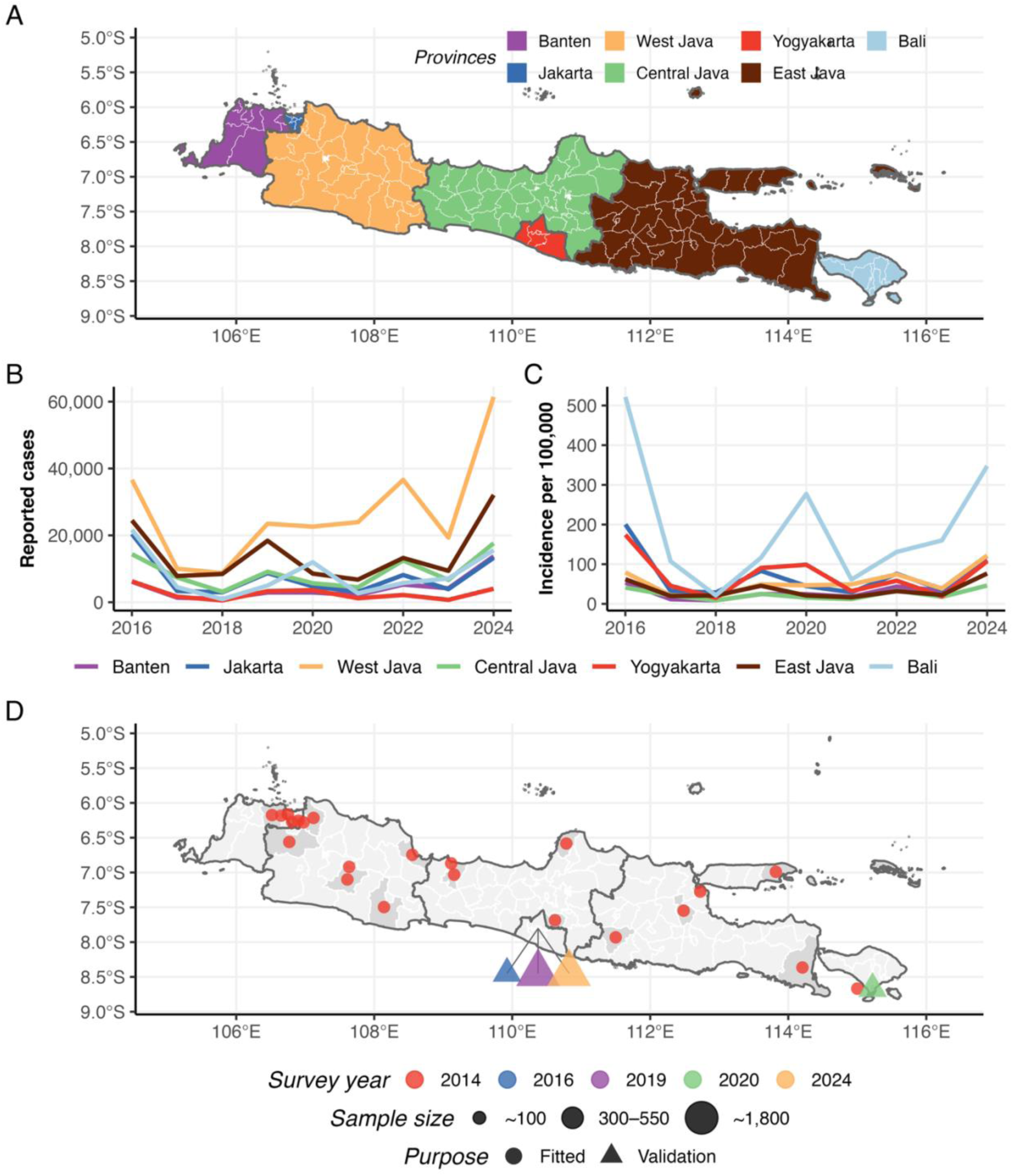
Study area and data overview. **(A)** Map of the study area comprising 7 provinces and 128 districts in Java and Bali, Indonesia. **(B)** Annual reported dengue haemorrhagic fever (DHF) cases by province, 2016–2024. **(C)** Annual incidence rate per 100,000 population by province. **(D)** Locations of seroprevalence surveys, coloured by survey year; circles indicate the survey fitted in the model (2014) and triangles indicate surveys held out for model validation. Point size indicates sample size.

#### Baseline FOI and reporting rate across model scenarios

The estimated national baseline FOI trajectory (2016–2024) captured the major inter-annual peaks across all four model scenarios (**Fig. 4A**). Models with covariates produced a notably higher baseline FOI (the national temporal component before covariate adjustment) in 2022 compared to models without covariates. In the covariate model, the Oceanic Niño index (ONI) term accounts for El-Niño-driven transmission in 2016 and 2024 (July to June mean ONI were 1.80 and 1.35, respectively, indicating moderate-to-strong El-Niño period), so the baseline need not rise as high in those years. In 2022, a non-El-Niño year, the baseline must absorb the full outbreak signal from case data, suggesting additional drivers of transmission beyond El Niño-Southern Oscillation variability. The estimated baseline reporting rate was the lowest in the full model (2.6%, 90% CrI: 2.1-3.1%) and the highest in case-data-only model (3.4%, 90% CrI: 2.8-4.1%) (**Fig. 4B**).

**Figure 4.**
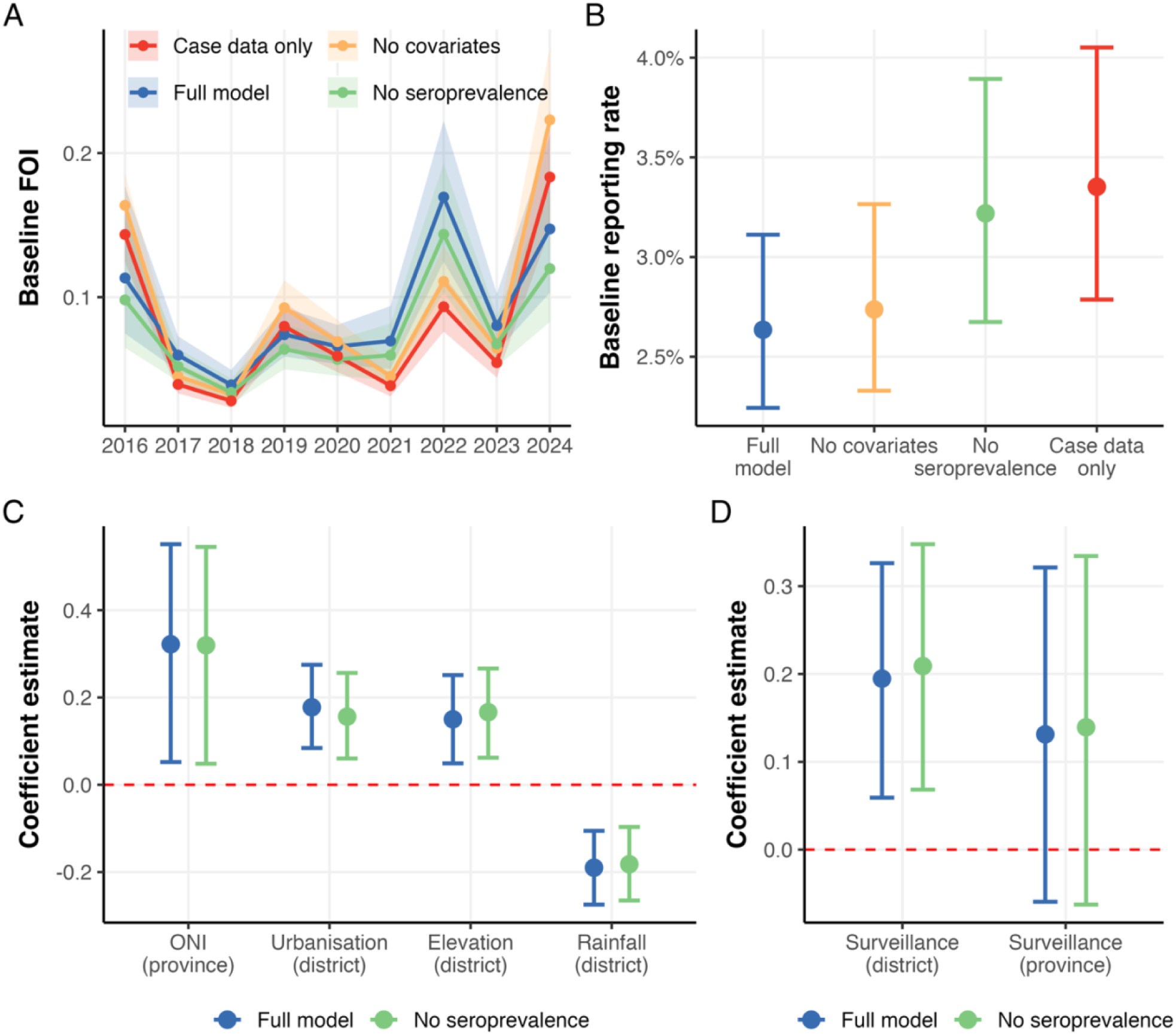
National baseline estimates and covariate effects across model scenarios. **(A)** Baseline total FOI (summed over the four serotypes), 2016–2024, for four model scenarios (posterior median with 90% CrI). **(B)** Baseline reporting rate (posterior median with 90% CrI) by model scenario. **(C)** Covariate coefficient estimates for FOI; points show posterior median, bars show 90% CrI. **(D)** Covariate coefficient estimates for reporting rate. Panels C–D compare the full model and the no-seroprevalence model (the two scenarios that include covariates).

Covariate effects on FOI were consistent between the full model and the no-seroprevalence (or covariates-only) model (**Fig. 4C**). Higher-than-average rainfall was associated with lower FOI, while ONI had the largest positive effect, though with the widest credible intervals. Urbanisation and elevation are both positively associated with FOI. The directions of the rainfall and elevation effects contrast with conventional expectations in dengue epidemiology and may reflect the specific ecological context of Java (see **Discussion**). District-level surveillance capacity was positively associated with reporting rate, while the province-level effect was not clearly distinguishable from zero (**Fig. 4D**).

#### Spatial and temporal FOI patterns

District-level FOI trajectories showed substantial heterogeneity both within and between provinces (**Fig. 5A**). Province-level median trajectories were broadly similar in magnitude and followed the national baseline pattern, with most provinces recording their highest estimated FOI during the study period in 2024 (though this may partly reflect improving surveillance absorbed into FOI estimates; see **Discussion**). However, individual districts showed considerable variation in both the timing and magnitude of transmission peaks, reflecting heterogeneous local transmission dynamics. The spatial distribution of mean FOI over the study period showed geographic clustering, with the highest FOI districts concentrated in the central part of West Java and in the southern and eastern parts of East Java (**Fig. 5B**). Lower FOI was estimated across most of Central Java and Yogyakarta.

**Figure 5.**
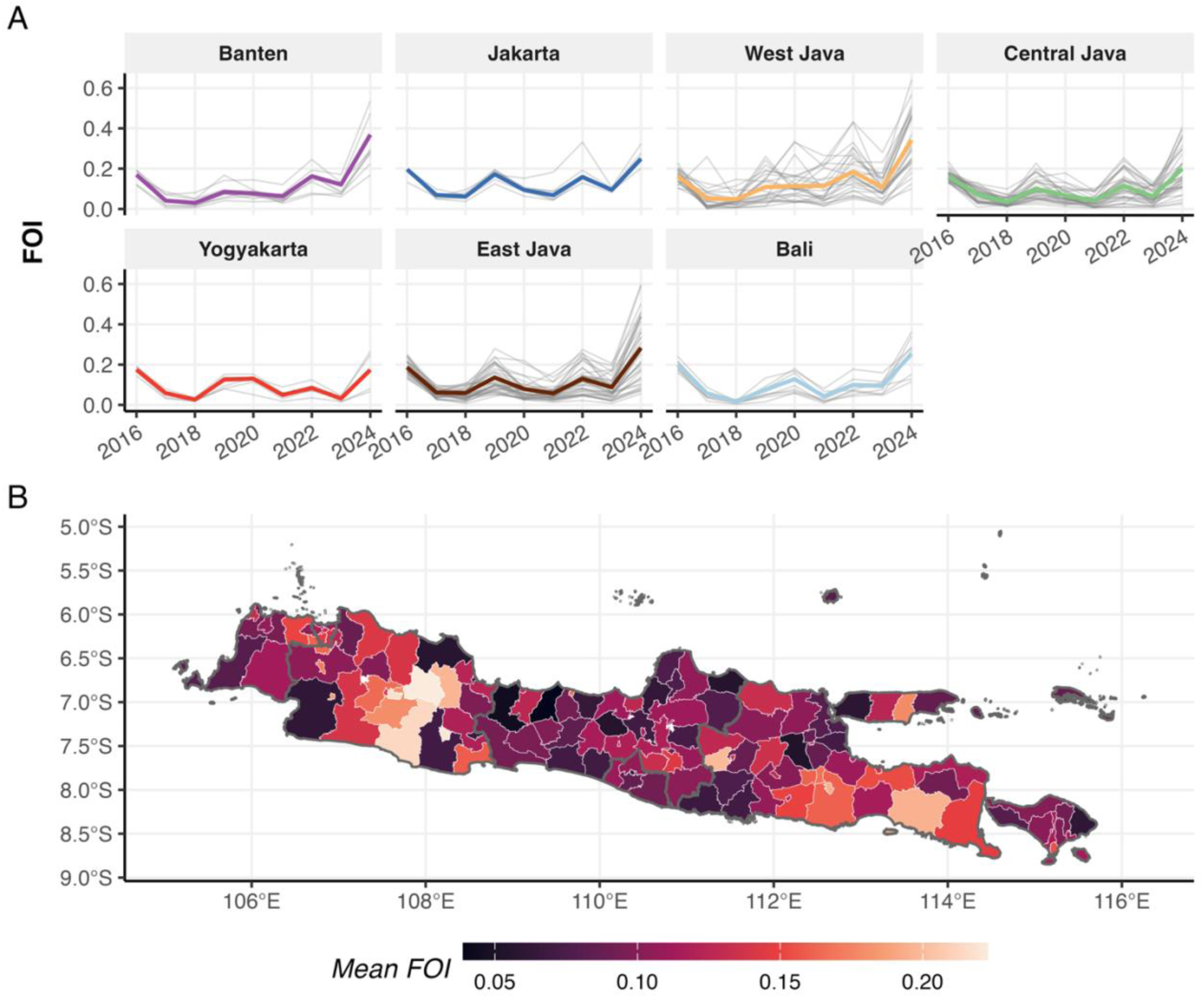
Spatial and temporal heterogeneity in total FOI (summed over the four serotypes) across Java-Bali, 2016–2024. **(A)** FOI trajectories by province; grey lines show individual district estimates (posterior medians), coloured lines show province-level medians. **(B)** Spatial distribution of mean FOI by district, averaged over the study period.

#### Reporting rate and surveillance heterogeneity

Estimated district-level reporting rates were spatially heterogeneous, with pockets of districts with higher reporting (**Fig. 6A**). At the province level, Jakarta and Bali had the highest median reporting rate (∼6.6% and ∼5.6%, respectively), followed by Yogyakarta (∼4.3%), while Banten, West Java, Central Java, and East Java had the lowest (∼2%) (**Fig. 6B**). At the district level, estimates ranged from 0.35% (90% CrI: 0.21-0.63%) in Wonosobo, Central Java, to 9.4% (90% CrI: 7.0-12.9%) in Kota Cirebon, West Java. The relative reporting ratio for primary versus secondary infection (*γ*) was estimated at approximately 0.2 across most provinces, consistent with prior expectation, although district-level estimates in Central Java varied more widely. Bali was a striking exception, with *γ* estimated at approximately 0.65, suggesting that primary infections contribute substantially to reported cases in that province (**Fig. 6C**). However, Bali contributes only aggregated case data, meaning that *γ* estimates in Bali are primarily informed by the hierarchical structure and seroprevalence data rather than age-specific case patterns, and should therefore be interpreted with caution. Surveillance capacity was positively associated with reporting rate at the district level in several provinces, though this relationship was weak, absent, or negative in Banten, Central Java, and East Java, provinces with persistently low reporting overall.

**Figure 6.**
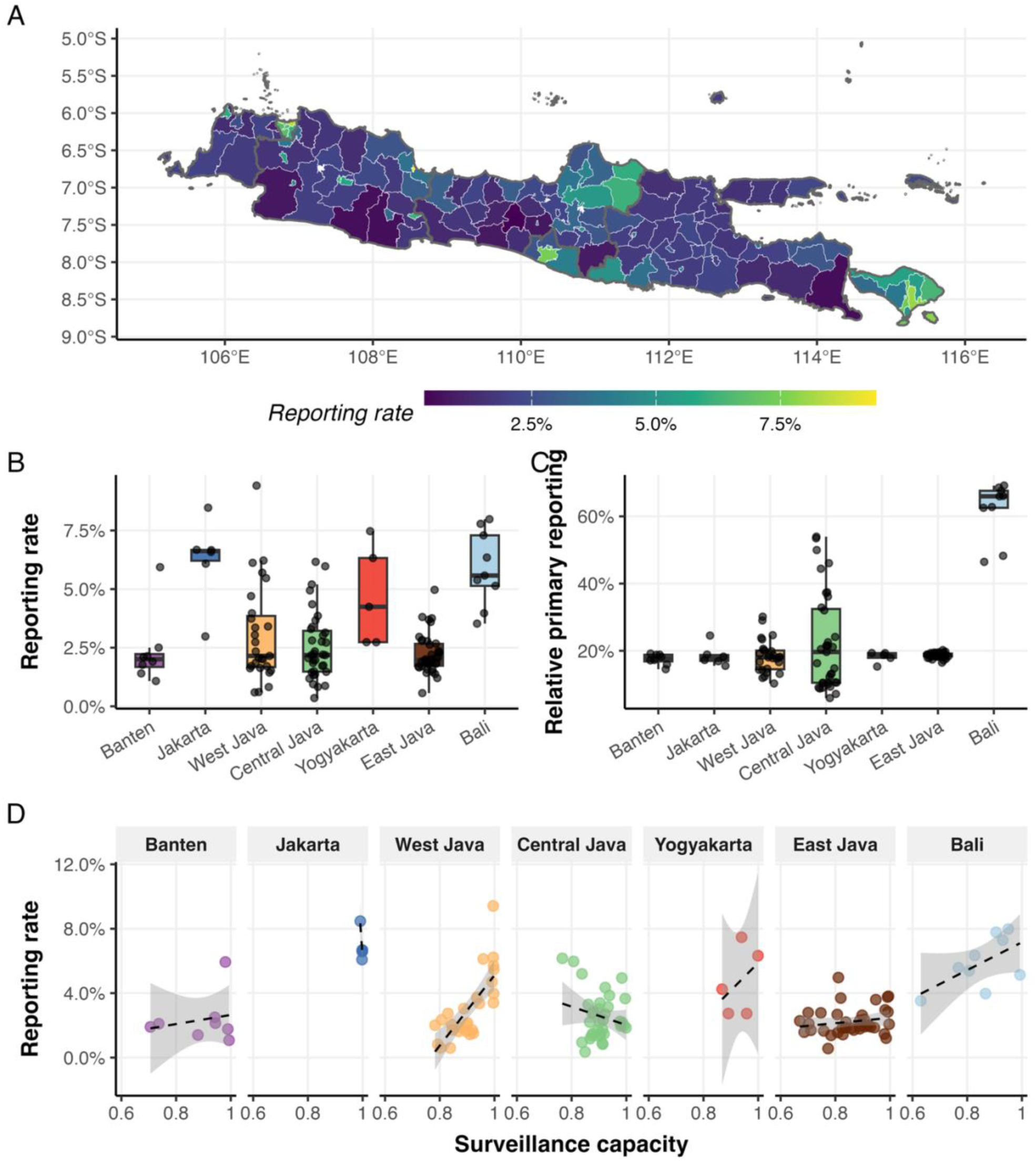
Reporting rate heterogeneity across Java-Bali. **(A)** Spatial distribution of district-level reporting rate (*ρ*, posterior median). **(B)** Distribution of reporting rates (*ρ*) by province (boxplots show median and interquartile range; points show individual districts). **(C)** Distribution of relative primary reporting ratio (*γ*) by province. **(D)** Estimated reporting rate versus district-level surveillance capacity, by province; dashed lines show within-province linear trends with 95% confidence bands.

#### Seroprevalence and vaccine eligibility

Predicted seroprevalence showed reasonable agreement with observed values across survey sites (R^2^=0.61, RMSE=0.20), with greater discrepancies among younger age groups for whom sample sizes were generally smaller (**Fig. 7A**; age-seroprevalence curves for each site in **Fig. S12**). Against held-out surveys, the model underpredicted the 2016 and 2019 surveys while closely matching the 2024 survey in Kota Yogyakarta, and slightly over-predicted seroprevalence among adults in Kota Denpasar (**Fig. 7B**).

**Figure 7.**
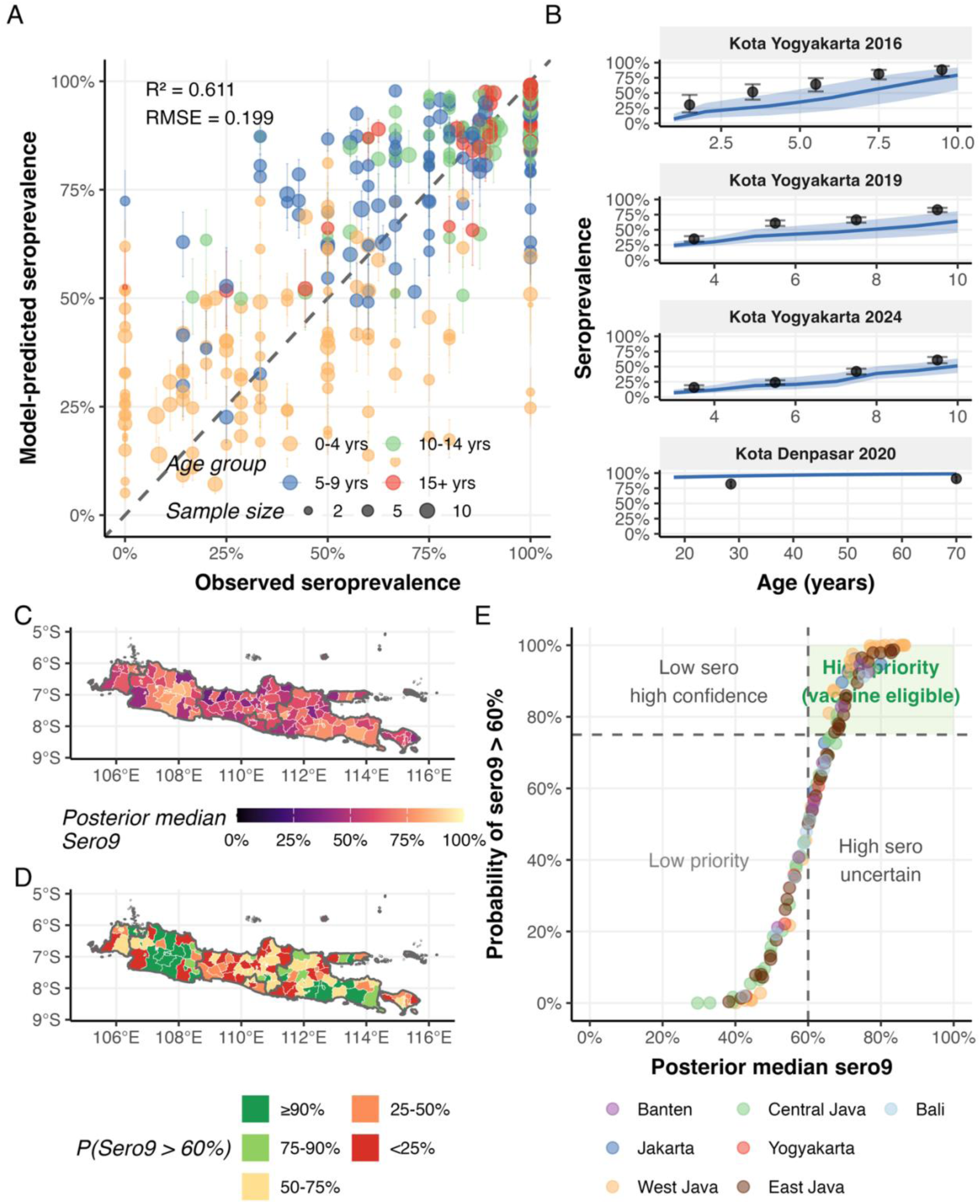
Seroprevalence estimates and vaccine eligibility classification. **(A)** Predicted versus observed seroprevalence by age group for the fitted 2014 survey sites (R² = 0.61, RMSE = 0.20); bars show 90% CrI, point colour indicates age group, and size indicates sample size; the dashed line indicates perfect agreement. **(B)** Age-seroprevalence curves for held-out surveys in Kota Yogyakarta (2016, 2019 and 2024) and Kota Denpasar (2020); blue line and shading show posterior median and 90% CrI, points and error bars show observed seroprevalence with 95% binomial confidence intervals. **(C)** Median estimated seroprevalence at age 9 by district at the end of 2024. **(D)** Posterior probability that seroprevalence at age 9 exceeds 60%, the WHO-suggested threshold for high dengue transmission intensity. **(E)** Policy prioritisation quadrant: median seroprevalence at age 9 versus posterior probability of exceeding 60%; dashed lines indicate classification thresholds (60% seroprevalence, 75% probability). Districts in the upper-right quadrant are classified as high-priority candidates for vaccine introduction.

Estimated median seroprevalence at age 9 exceeded 60% in 79 of 128 districts by the end of 2024, with the highest values in Jakarta, West Java and East Java, while districts in Central Java, Yogyakarta and Bali were spread around the threshold (**Fig. 7C**). Accounting for posterior uncertainty, 49 districts had at least a 75% posterior probability of exceeding 60%, concentrated in West Java and East Java (**Fig. 7D**). The policy prioritisation quadrant (**Fig. 7E**) identified these 49 districts as high-priority candidates for vaccine introduction, while a further 30 districts had median seroprevalence above 60% but with ‘insufficient’ certainty. In Bali, only Kota Denpasar was classified as high priority, despite the province having the highest reported incidence among study provinces.

#### Misalignment between model-based and incidence-based vaccine prioritisation

We compared the model-based vaccine eligibility classification (requiring both a posterior median seroprevalence at age 9 above 60% and a posterior probability of exceeding 60% greater than 75%) against an incidence-based criterion that selected the same number of districts with the highest mean reported incidence rate over 2016-2024 (a simple approach that might be used in the absence of seroprevalence data or modelled estimates). Of the 49 districts identified as high priority by each approach, only 23 were concordant, close to the 19 expected if districts had been selected at random (49 districts with each 49/128 chance of matching model-identified districts) (**Fig. 8A**).

**Figure 8.**
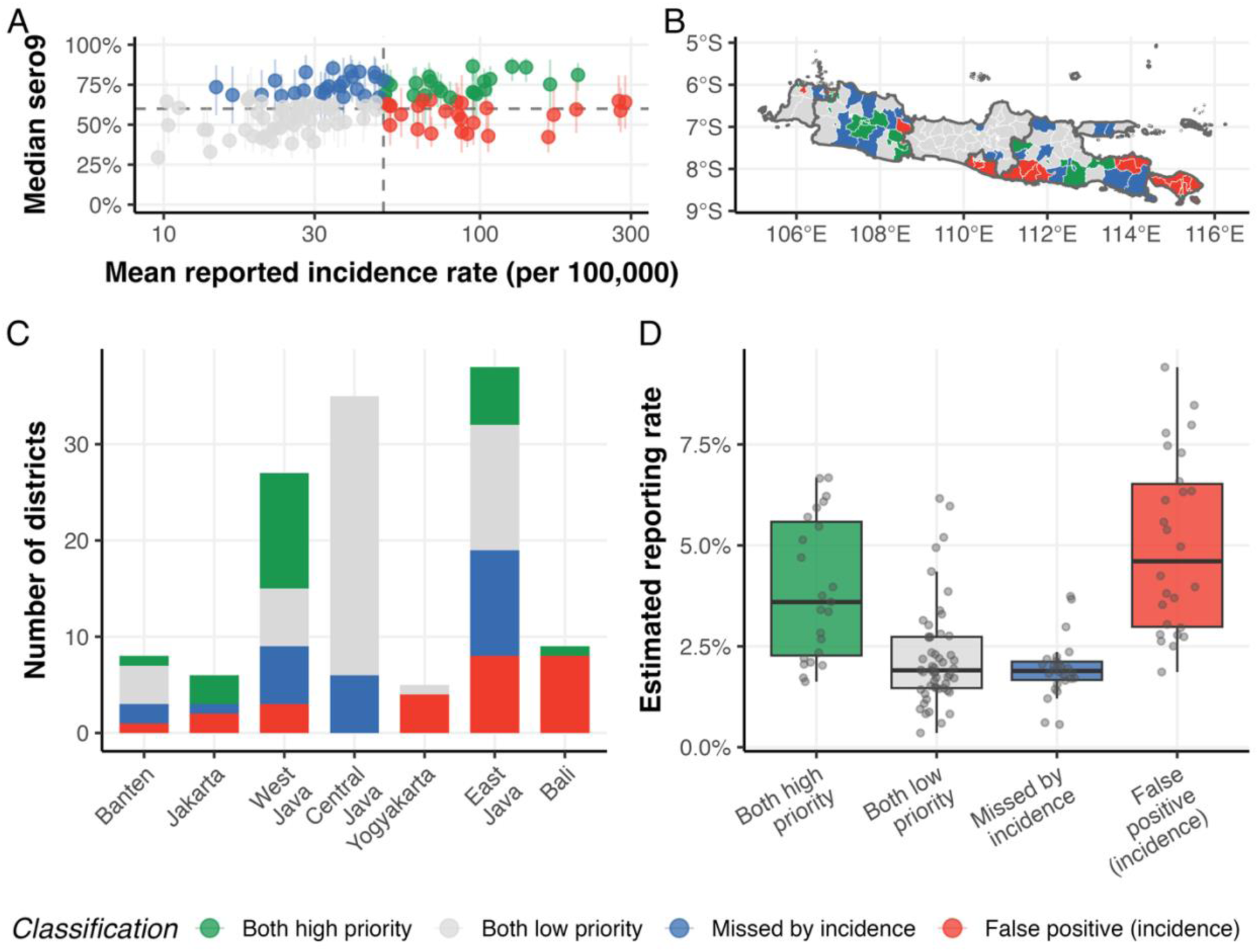
Misalignment between model-based and incidence-based vaccine prioritisation. **(A)** Median seroprevalence at age 9 (with 90% CrI) versus mean annual reported incidence rate (per 100,000, log scale) by district; dashed lines indicate classification thresholds (60% seroprevalence; incidence cut-off selecting the same number of districts as the model, n = 49). **(B)** Spatial distribution of agreement categories across Java and Bali. **(C)** Number of districts in each agreement category by province. **(D)** Estimated reporting rate (posterior median) by agreement category.

Geographic agreement between the two approaches is shown in **Fig. 8B–C**. Districts classified as high-priority by modelled seroprevalence but missed by the incidence-based approach were concentrated in West Java, Central Java and East Java, while districts flagged by incidence but not by the model were concentrated in Bali, Yogyakarta and also East Java. Grouping the reporting rates of each district by agreement between model-based and incidence-based approaches (**Fig. 8D**), districts identified as high-priority by the model missed by the incidence-based approach had lower estimated reporting rates than concordant high-priority districts, similar to districts classified as low-priority by both, while false positive districts had the highest reporting rates. A sensitivity analysis using more lenient thresholds (median seroprevalence at age 9 > 60% versus the same number of top-incidence rate districts) showed a similar pattern, with 58 of 79 high-priority districts concordant between the two approaches, compared with 49 expected by chance (**Fig. S13**).

#### Model comparison

Leave-one-out cross-validation (LOO-CV) on the case data showed no meaningful differences between models with and without covariates, whereas two models fitted to seroprevalence data had a lower expected log pointwise predictive density (ELPD) than those fitted to case data alone (**Table S1**). On the seroprevalence data, and on the joint case and seroprevalence likelihood, the full and no-covariate models performed equivalently. Including the 2014 serosurvey therefore came at a small cost in fit to the case data, suggesting some tension between age profile of reported cases and the population immunity implied by the survey, while improving agreement with both the fitted and held-out serosurveys (**Fig. S14**). Although around 20% of case observations had Pareto k values above 0.7, the difference between models with and without seroprevalence data was not driven by these observations (**Table S1**). Accounting for all these, we consider the full model the most suitable for application in this and similar settings, as it integrates all available data sources, gave the lowest prediction error for the fitted and held-out serosurveys.

## DISCUSSION

We present a Bayesian hierarchical catalytic model that jointly estimates dengue FOI from age-stratified case data, aggregate case data, and seroprevalence surveys within a single inference framework. This extends previous catalytic modelling approaches (*7–12*), which typically require age-stratified data from all locations, by allowing districts with different levels of data completeness (and different age grouping format) to contribute simultaneously through a hierarchical structure and shared covariate effects.

Synthetic validation demonstrated robust parameter recovery across four data completeness scenarios, even when covariates explained only small share of between-district variation in FOI. Age-stratified case data are a primary source of information for distinguishing FOI and reporting rates, since the age pattern of cases reflects cumulative exposure history and relative contribution of primary and secondary infections. Consistent with this, reporting rates were well recovered whenever age-stratified data were available, with little loss when only four age groups were reported, but were underestimated in the scenario where most districts reported aggregate data only (S4). In that setting, district reporting rates were shrunk towards the shared mean. FOI and seroprevalence, in contrast, remained reasonably well estimated in S4, with only moderately higher error than in scenarios with full age stratification. Covariates contributed little to the accuracy of point estimates but improved the calibration of credible intervals. These findings have practical implications for dengue surveillance in low- and middle-income countries that do not routinely report age-stratified case data. Age-stratified reporting from a subset of locations, even with coarse age groups, can support FOI and seroprevalence estimation across a wider network of districts reporting only aggregate totals, whereas reporting rate estimates for aggregate-only districts are better interpreted in relative terms, as an indication of which districts report more or less completely, than as absolute terms.

Adding seroprevalence data to the synthetic data fits mainly improved seroprevalence estimates for cohorts whose exposure depended on historical FOI, with little effect on estimates at the end of the surveillance period (**Figs. S3-S7**). In the real data, however, the serosurvey also improved predictions of held-out surveys conducted after the surveillance period began, most visibly in Kota Yogyakarta in 2024 (**Fig. S14**). This contrast likely reflects that the synthetic data were generated with a constant reporting rate, so case data alone could constrain recent transmission, whereas in reality, reporting may change over time with improvements or disruptions in surveillance. In the real data, fitting the 2014 serosurvey also slightly worsened the fit to case data, suggesting tension between the immunity implied by the survey and case patterns that a single reporting rate per district cannot fully accommodate. Because serosurveys measure population immunity directly, independent of reporting, periodic serosurveys could help anchor estimates of population immunity, particularly in settings where surveillance performance changes over time. We further note that our synthetic data were generated under the same model structure that was then fitted, so these experiments test identifiability under a correctly specified model, and the performance reported here should be regarded as an upper bound. Our misspecification checks provide some reassurance, as an uninformative covariate was correctly estimated to have no effect, and including only a subset of the true covariates gave performance between that of the full- and no-covariate models (**Figs. S8-S11**).

A well-known challenge in catalytic models is the identifiability of FOI and reporting rate from case data alone, as many combinations of these parameters can produce similar expected case counts. In our framework, this trade-off was resolved mainly by the age pattern of reported cases, which reflects cumulative exposure, and, for transmission history, by seroprevalence data. The contribution of covariates depended on how much of the between-district variation in FOI they explained. In the setting most comparable to Java and Bali, where covariates explained around 10% of this variation, they mainly improved the calibration of uncertainty rather than the accuracy of point estimates. Some of the individual coefficients were weakly identified, with some biased in a consistent direction across scenarios. As covariates explained a larger share of variation, FOI accuracy improved substantially and coefficient bias largely disappeared (**Fig. S1**), indicating that covariates become increasingly valuable where transmission is more strongly shaped by measurable drivers. In the real data, covariates explained a small share of variation in FOI, so the estimated covariate effects (**Fig. 4C**) are best interpreted as associations that help the model to estimate FOI across districts, rather than as precise measures of the effect of each driver.

Applied to nine years of surveillance data across Java and Bali, the model revealed a substantial spatial heterogeneity in both FOI and reporting rates at the district level. This within-province variation underscores the importance of subnational, district-level modelling for dengue policy decisions.

The national baseline reporting rate (2.6-3.4% across model scenarios) was somewhat lower than expert panel estimates of around 5% (*15*), although absolute reporting rates depend on model assumptions and are better interpreted in relative terms across districts. Bali stood out as one of the provinces with the highest reporting rates and a strikingly high relative primary reporting ratio (*γ*=0.65, compared with ∼0.2 elsewhere). Bali’s healthcare infrastructure, oriented towards a large international and local tourist population, may facilitate detection of milder dengue presentations. Travelers who seek care locally but are absent from the population denominator would inflate both estimated reporting rate and relative reporting ratio, as the model needs to accommodate excess cases not predicted by the resident population structure. Alternatively, if FOI in Bali is indeed lower than elsewhere, primary infections would occur at older ages, when infections are more likely to be symptomatic, prompting healthcare-seeking (*16*). The model also slightly overestimated seroprevalence in Bali relative to survey data, including among adults in the held-out Kota Denpasar survey (**Fig. 7B**), which could reflect genuinely lower historical FOI than estimated, or the absence of waning immunity in the model. While the explanations above are plausible, we note that Bali’s *γ* estimates are less well constrained than in provinces with age-stratified data, and the high value could partly reflect identifiability limitations.

When appropriate covariates are included, the model can partially disentangle climate-driven and non-climate-driven components of inter-annual variation. The covariate model attributed the 2016 and 2024 outbreaks partly to El Niño conditions (12-month mean ONI of 1.80 and 1.35, respectively), while producing a notably higher baseline FOI in 2022, a non-El Niño year (*17*). This residual signal may reflect unmeasured drivers such as serotype replacement, though direct evidence is limited. A shift toward DENV-3 was documented in Bali during this period (*18*), which has been associated with outbreak dynamics elsewhere in Southeast Asia (*19*). However, whether similar shifts occurred across Java remains unknown due to limited serotype surveillance. Additionally, our model assumes equal FOI across serotypes and cannot directly capture serotype-specific dynamics. The gradual return of population mobility following relaxation of COVID-19 restrictions may also have some influence during this period (*20*, *21*).

The negative association between above-average rainfall and FOI, and the positive association between elevation and FOI, both contrast with conventional expectations in dengue epidemiology, where rainfall is generally assumed to promote *Aedes* breeding and higher elevation to limit transmission through lower temperatures (*22*). These associations may reflect the specific context of our study region. Above-average rainfall in an already wet environment may flush larval breeding sites (*23*, *24*). Similarly, the positive elevation effect may reflect the fact that many of Java’s densely populated urban centres are located at higher elevations, and that the temperature range across Java’s populated areas, even at those with higher elevations, remains within the suitable range for *Aedes* survival and dengue transmission (*25*). These findings highlight that covariate effects estimated in one ecological setting should not be directly extrapolated to other contexts without consideration of local geography.

The year 2024 recorded the highest estimated FOI across most provinces during the study period. While this may reflect a genuine increase in transmission, the assumption of constant reporting rate over time means that temporal improvements in surveillance capacity will be absorbed into FOI estimates, potentially inflating recent values. Our synthetic validation also showed that seroprevalence estimates become less accurate towards the end of surveillance period, as FOI in the final years is less constrained (**Fig. 2A**). Allowing time-varying reporting in future model extensions could help distinguish genuine transmission increases from improved case detection or changes in surveillance criteria, though this would introduce additional identifiability challenges that would likely require informative covariates or external data, such as periodic serosurveys, to resolve.

A key application of this framework is informing subnational vaccine targeting. The WHO 2024 position paper recommends the use of TAK-003 (Qdenga) in settings with high dengue transmission intensity, for which seroprevalence at age 9 exceeding 60% has been suggested as an indicator (*5*). Our estimates indicate that more than half of districts in Java and Bali (79 of 128) exceed this threshold, but applying a more conservative criterion requiring posterior probability greater than 75% reduces this number to 49. This highlights the importance of propagating uncertainty into policy decisions rather than relying on point estimates alone. Although estimates towards the end of the surveillance period are less accurate, seroprevalence at age 9 in 2024 remained well calibrated in synthetic validation and the model closely matched the held-out 2024 serosurvey in Kota Yogyakarta (**Figs. 2B, 7B**). In practice, vaccination decisions may be made at province or national level, and district-level estimates could be aggregated using population-weighted averages to inform decisions at broader spatial scales.

The comparison between model-based seroprevalence classification and incidence-based prioritisation revealed substantial misalignment. When uncertainty in seroprevalence estimates was accounted for, an incidence-based selection of the same size performed just slightly better than a random selection in identifying high-priority districts. Districts identified as high-priority by seroprevalence but missed by incidence were concentrated in West Java, Central Java and East Java, provinces where most districts had low estimated reporting rates. This suggests that passive surveillance underestimates transmission intensity where case detection is less complete. Conversely, districts flagged by incidence but not by the model included nearly all districts in Bali and Yogyakarta, where higher estimated reporting rates inflate reported case counts relative to the underlying seroprevalence. This pattern suggests that vaccine allocation based solely on reported incidence would systematically under-serve areas with the greatest surveillance gaps while over-prioritising areas with better case detection.

Model-based seroprevalence estimation offers a more consistent basis for identifying high-transmission districts, as it integrates multiple data sources and is less susceptible to surveillance bias than incidence-based approaches. However, this approach requires investment in both data infrastructure, particularly seroprevalence surveys, covariate data and age-stratified surveillance data, as well as analytical capacity and computing infrastructure. Seroprevalence surveys, even at limited spatial coverage, are particularly valuable as they constrain population immunity independently of reporting. In this study, we fitted only the 2014 serosurvey and retained others for validation, but in practice, all available serosurveys should be included to strengthen estimation.

Several limitations should be noted. First, the model assumes a constant reporting rate over the study period. As discussed above, temporal changes in surveillance capacity or case definition would be absorbed into FOI estimates, potentially biasing recent estimates upwards. Second, the model assumes equal FOI across the four dengue serotypes and does not capture serotype-specific immunity or replacement dynamics. In hyperendemic settings where serotype composition fluctuates, this simplification may obscure important drivers of inter-annual variation. Third, the model does not incorporate waning of dengue antibodies, which may lead to overestimation of seroprevalence, particularly among adults. Fourth, while the covariate framework improves parameter identifiability, substantial uncertainty in district-level estimates remains, particularly for districts with aggregate-only case data, where reporting rate tend to be shrunk towards the shared mean. Expanding routine age-stratified case reporting to these districts would provide the information needed to separate transmission intensity from reporting and would improve the precision of future estimates. Fifth, the temporal structure is simplified, as historical FOI uses a constant level for the distant past and district deviations from the national baseline are fixed near zero in the first year of data, so that between-district differences in that year are attributed mostly to covariates, which may bias their coefficients. Two structural choices also remain untested. Provinces serve as the intermediate grouping unit in the hierarchy, but alternative groupings, such as ecological zones or urban-rural clusters, could yield different pooling behaviour, and we did not assess sensitivity to this choice. Similarly, the AR(2) specification for district-level temporal residuals was selected *a priori*, and more constrained or more flexible temporal structures may be preferable in settings with longer or shorter surveillance series.

Several directions for future work emerge from this study. Allowing time-varying reporting rates, informed by external data on surveillance system changes, would improve interpretation of temporal FOI trends. Incorporating serotype-specific dynamics, potentially informed by serotype surveillance data, would enable the model to capture serotype replacement as a driver of outbreak variation. Expanding the geographic scope beyond Java and Bali to other Indonesian islands, and to other dengue-endemic countries with heterogeneous surveillance systems, would further test the generalisability of the framework.

In conclusion, our framework enables district-level estimation of dengue transmission and population immunity across a heterogeneous surveillance landscape, in which districts contribute different combinations of age-stratified and aggregate case data, with varying age groupings and formats that may change over time, supplemented by serosurveys where available. Applied to Java and Bali, these estimates showed that reported incidence is a poor guide to where population immunity is high, because differences in case detection capacity between districts bias incidence-based rankings. Such modelling complements rather than replaces improvements in surveillance. Expanding the collection of age-stratified case data remains crucial, and together with periodic serosurveys, they would provide a stronger basis for monitoring transmission and for targeting vaccination where it is most needed.

## MATERIALS AND METHODS

We developed a Bayesian hierarchical catalytic model to estimate time-varying dengue force-of-infection (FOI) across multiple districts and provinces simultaneously, integrating routine surveillance data and seroprevalence surveys within the parameter inference framework. The model extends previous catalytic modelling approaches (*10*) by accommodating multiple provinces and districts within a hierarchical structure, incorporating external covariates to inform FOI dynamics and reporting rate, allowing districts to contribute data in different formats (age-stratified or yearly aggregate only), and jointly fitting to both incidence and seroprevalence data, leveraging complementary information regarding transmission history. The youngest age included in both seroprevalence and case data was 1 year (i.e., children who have completed their first year of life), by which time maternally transferred antibodies are expected to have waned.

### Study areas and data sources

#### Study settings

We analysed dengue transmission dynamics across seven provinces in Java and Bali, Indonesia, comprising 128 districts over the period of 2016–2024. Java and Bali together account for approximately 60% of Indonesia’s population and consistently report the highest dengue burden nationally. The study area spans a wide range of urbanisation levels, population densities, and ecological conditions, providing a suitable setting for examining spatial heterogeneity in dengue transmission and reporting.

#### Dengue surveillance data

District-level dengue case data were obtained from the routine surveillance system of the Arbovirus Control Programme of the Indonesia Ministry of Health. Indonesia’s national surveillance system reports clinically diagnosed dengue haemorrhagic fever (DHF) cases. All seven provinces contributed to nine years of surveillance data (2016–2024).

The availability and format of case data varied across provinces and over time (**Table 1**). Some provinces provided age-stratified case counts for most years, with Jakarta reporting nine age groups (1–4, 5–9, 10–14, 15–19, 20–44, 45–54, 55–64, 65–74, >74), and other provinces reporting only four age groups (1–4, 5–14, 15–44, >44). Two provinces (East Java and Bali) contributed only aggregate annual case totals for the entire study period, while several provinces had a mix of years with and without age stratification.

#### Population data

District-level population data were obtained from linear interpolations and extrapolations of the 2010 and 2020 census data (*26*, *27*). Interpolations were done by sex and age group for each district.

#### Seroprevalence surveys

We incorporated data from five seroprevalence surveys of dengue in Indonesia. The nationwide urban serosurvey was used for model fitting, and the remaining were held out for out-of-sample validation:

1. <u>Nationwide urban serosurvey</u> (*28*). This cross-sectional survey was conducted in 2014 across 30 urban sites in Indonesia, including sites within six provinces (all except Yogyakarta) in the study area. Participants aged 1–18 years were tested for anti-dengue IgG antibodies. A total of 2,342 individuals from 22 sites within our study area were included.
2. <u>Pre-wolbachia trial serosurvey in Yogyakarta</u> (*29*). Baseline seroprevalence data were collected in 2016 prior to the Wolbachia trial in Yogyakarta. Participants aged 1–10 years from Yogyakarta City were tested for IgG antibodies. 314 individuals were included.
3. <u>Wolbachia trial and post-deployment serosurveys in Yogyakarta</u> (*30*). Children aged 3-10 years in Kota Yogyakarta were tested for IgG antibodies during the cluster-randomised trial in 2018-2019 (1,727 individuals) and after the city-wide Wolbachia deployment in 2023-2024 (1,821 individuals).
4. <u>Bali serosurvey</u> (*31*). A seroprevalence survey was conducted in 2020 in Denpasar City of Bali province. Participants aged 18 years and older were tested for IgG antibodies. 539 individuals were included.

#### Covariates

We included environmental, climatic, and health system covariates to inform heterogeneity in both FOI and reporting rates across districts. Covariates were selected based on established associations with dengue transmission ecology and data availability in the study region. **Table 2** shows the covariates used for the province and district levels for each FOI and reporting rate model, including data sources of those covariates. Rainfall was included rather than temperature as previous analysis of this region found rainfall to be a stronger predictor of dengue transmission (*17*), consistent with Java’s narrow tropical temperature range being largely suitable for *Aedes* survival (*25*).

**Table 2.** Covariates included in the FOI and reporting rate models.

| **Model component** | **Covariate** | **Level** | **Temporal resolution** | **Source** | **Processing** |
|---|---|---|---|---|---|
| FOI | Oceanic Niño Index (ONI) | Province | Time-varying | (*32*) | As is, same values for all provinces; accounting for 6-month lag (July-June average) |
| FOI | Urbanisation | District | Constant | (*33*) | Population weighted |
| FOI | Mean elevation | District | Constant | (*34*) | Population weighted |
| FOI | Annual rainfall | District | Time-varying | (*35*) | Population weighted; accounting for 3-month lag |
| Reporting rate | Surveillance capacity | Province | Constant | (*4*) | Population weighted |
| Reporting rate | Surveillance capacity | District | Constant | (*4*) | Population weighted |

### Dengue hierarchical catalytic model

#### Epidemiological assumptions

The underlying epidemiological assumptions remain similar to previous catalytic modelling frameworks without serotype data (*7–12*). We assume four co-circulating dengue virus (DENV) serotypes with equal per-serotype FOI (*λ*). Individuals fully susceptible to all serotypes are exposed to a combined FOI of 4*λ*. Following the primary infection with one serotype, individuals enter a monotypically immune state, during which they are immune to the infecting serotype but remain susceptible to the remaining three serotypes (exposed to 3*λ*). Secondary infections are more likely to cause severe disease due to antibody-dependent enhancement (ADE) and therefore more likely to be detected by the surveillance system than primary infections. We assume tertiary and quaternary infections contribute negligibly to reported incidence.

#### Hierarchical structure

The model employs a two-level hierarchical structure. Provinces (*p* = 1, 2, …, *n_P_*) serve as intermediate grouping units, each containing multiple districts (*d* = 1, 2, …, *n_Dtotal_*), with district *d* nested within province *p*. All districts contribute data over the same time interval (*n_Ttotal_* years), ensuring temporal alignment of the baseline FOI across locations.

#### Infection dynamics

For each district *d*, we track the proportion of the population by age *a* (in years) and year *t* in two immunity states: fully susceptible to all four serotypes, *S*(*a*, *d*, *t*), and monotypically immune, *M*(*a*, *d*, *t*). Fully susceptible individuals are exposed to all four serotypes at a combined rate of 4*λ_d_*_,*t*_, while monotypically immune individuals are exposed to the three remaining serotypes at a combined rate of 3*λ_d_*_,*t*_, where *λ_d_*_,*t*_ is the per-serotype FOI for district *d* in year *t*. The annual incidence of primary (*I*_1_) and secondary infections (*I*_2_) for individuals age a in district *d* during year *t* are:

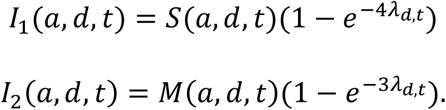

These are exact annual infection probabilities given the per-year hazard, and each individual can experience at most one infection per year, reflecting short-term cross-protection following infection (*36*). Accounting for infections during the year and aging of each birth cohort, the proportions susceptible and monotypically immune at the start of the following year are:

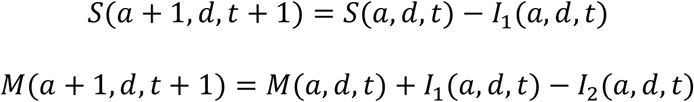

Individuals are born fully susceptible, i.e., *S*(0, *d*, *t*) = 1 and *M*(0, *d*, *t*) = 0.

#### Reporting model

Secondary dengue infections typically present with more severe clinical manifestations due to ADE and are thus more likely to be detected by the surveillance system. The expected number of reported cases in age group *g* for district *d* in year *t* is:

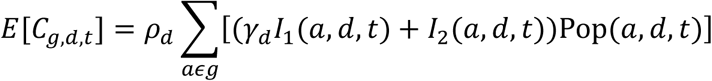

where Pop(*a*, *d*, *t*) is the population aged *a* years in district *d* at time *t*, so that expected cases are summed over the single-year ages in each age group. Single-year populations were obtained by dividing each 5-year census age band equally across its single years of age, up to a maximum age of 99 years. Here, *ρ_d_* is the district-level reporting rate for secondary infections, *γ_d_* is the relative reporting ratio for primary versus secondary infections (with *γ_d_* < 1 expected in endemic settings), and *Pop_g_*_,*d*,*t*_ is the population in age group *g*. The summation is taken over all single-year ages *a* within the age group *g*. The total expected cases across all age groups is:

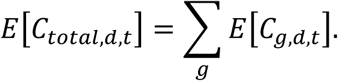

The specification and estimation of *ρ_d_* and *γ_d_* are described in the ‘Reporting rate model’ section.

#### FOI model

##### FOI specification

The time-varying per-serotype FOI (*λ_d_*_,*t*_) for district *d* in year *t* is modelled on the logit scale as:

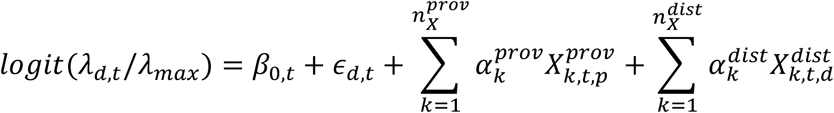

where *β*_0,*t*_ is a national time-varying baseline FOI, *ε_d_*_,*t*_ is a district-specific temporal residual following an AR(2) process, 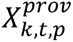 and 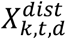 are province- and district-level covariates with coefficients 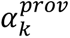 and 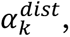, respectively (**Table 2**), and *λ_max_* = 0.25 is an upper bound reflecting the maximum plausible annual infection risk per serotype, which corresponds to a maximum total FOI of 1.0 across all four serotypes. Empirical estimates of total FOI in hyperendemic settings rarely exceed 0.25 (*8*, *9*), so this bound is not restrictive and the formulation allows FOI to vary annually while borrowing information across districts through the shared baseline and covariate effects. Posterior draws confirm that this bound is not materially restrictive: across all district-years the largest posterior median was 0.160, and the largest upper 90% credible bound was 0.208, or 83% of the bound, although the single largest posterior draw reached 0.244 (**Fig. S15**).

##### National time-varying baseline of FOI

The national baseline *β*_0,*t*_is modelled as a first-order random walk on the logit scale:

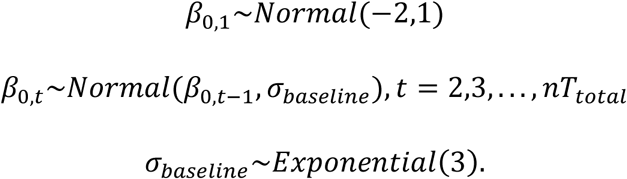

This formulation allows smooth temporal variation in the baseline transmission intensity while partially pooling information across years.

##### District-level temporal residuals: AR(2) process

To capture potential multi-year cycles in dengue transmission (e.g., relating to serotype dynamics or climate periodicity), district-specific residuals *ε_d_*_,*t*_ follow an autoregressive process of order 2:

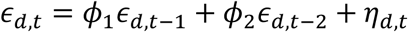

where *φ*_1_ and *φ*_2_ are shared autoregressive coefficients and *η_d_*_,*t*_∼*Normal*(0, *σ_ε_*) is white noise. For computational efficiency, we employ non-centred parameterisation for *t* ≥ 3, where the first two time points are sampled directly (centred), with the first-year residual constrained near zero, *ε_d_*_,1_∼*Normal*(0,0.01). The priors are:

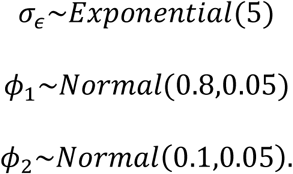

These priors favour first-order autocorrelation, reflecting the expected persistence of transmission intensity from year to year, with a smaller second-order component to capture potential multi-year cyclical patterns. Although these priors are informative, prior-posterior comparison indicates that the temporal parameters are informed by the data rather than prior-driven: the posterior for the first-order coefficient concentrated at 0.617 (90% CrI 0.564-0.667), some 3.7 prior standard deviation below the prior mean of 0.80, while the second-order coefficient remained close to its prior (posterior 0.098, 90% CrI 0.047-0.151) (**Fig. S16**).

##### Covariate effects on FOI

Province-level and district-level covariates (**Table 2**) are included to inform spatial heterogeneity in FOI across districts. Covariate regression coefficients have priors:

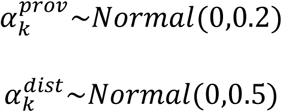

The prior variance is larger for district-level effects, reflecting greater uncertainty in local-scale covariate-transmission relationships. All covariates were standardised to mean zero and unit standard deviation prior to model fitting.

##### Historical FOI and population immunity initialisation

To initialise population immunity at the start of surveillance data availability, we estimate FOI during the historical period preceding the data. We use a two-phase approach, with a single district-specific shift applied to both phases.

###### Recent history (10 years before surveillance data)

Annual FOI follows a national trajectory, shifted up or down by a district-specific level:

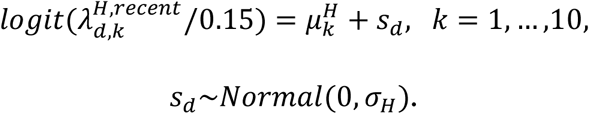

where *k* is the index for years before the surveillance data (*k* = 1 being the year immediately before), 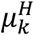 is the national recent-history FOI on the logit scale, and *s_d_* is the district shift with between-district standard deviation *σ_H_*, with priors:

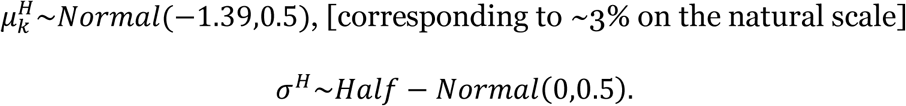

Historical FOI is bounded at 0.15 per serotype. Because case data inform only cumulative exposure of each cohort before surveillance, not its timing, a single shift per district is estimated rather than district-specific annual values.

###### Distant past (>10 years before surveillance data)

FOI is assumed constant over time at a national level, *λ^H^*^,*constant*^, shifted by the same district level *s_d_*:

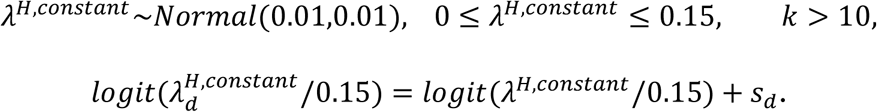

The prior is truncated at zero in the implementation, so that historical FOI is constrained to be strictly non-negative on the natural scale.

The 10-year cutoff provides sufficient temporal detail to inform immunity accumulation in cohorts aged 1–10 years at the start of surveillance, while avoiding over-parameterisation for older periods for which the data provide little constraint.

###### Immunity initialisation

Susceptibility and monotypic immunity at the first surveillance year are obtained by applying the annual update equations (see **Infection dynamics**) over the historical period. Each cohort aged at the start of surveillance is exposed to 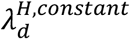 for its years of life more than 10 years before the data and to 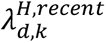 for the 10 years immediately before.

#### Reporting rate model

##### Reporting rate specification

dThe overall reporting rate *ρ_d_* for district *d*, defined as the proportion of all dengue infections detected and reported by the surveillance system, is modelled as:

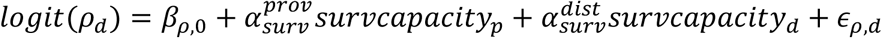

where *β_ρ_*_,0_ is the national baseline reporting rate, *survcapacity_p_* and *survcapacity_d_* are standardised surveillance capacity indices for province *p* and district *d*, respectively (derived from (*4*); **Table 2**), quantifying surveillance quality in capturing viral infections (from (*4*)), 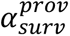 and 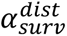 are corresponding regression coefficients, and *ε_ρ_*_,*d*_∼*Normal*(0, *σ_ρ_*) is a district-specific random effect.

The priors are:

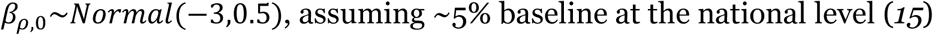

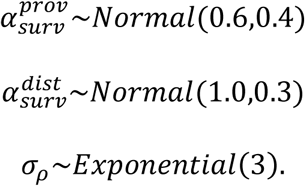

The informative prior on *β_ρ_*_,0_ reflects existing estimates of dengue reporting rates in Indonesia (*15*), while the positive-centred priors on surveillance capacity coefficients assume the expectation that higher surveillance capacity is associated with higher case detection. Because these priors are deliberately optimistic, we compared them against their posteriors to verify that the reported associations are not an artefact of the prior. The district-level coefficient shifted 2.7 prior standard deviations below its prior mean (posterior 0.195, 90% CrI 0.059-0.326, against a Normal(1.0, 0.3) prior), and the province-level coefficient was not distinguishable from zero (posterior 0.131, 90% CrI -0.059-0.321). The data therefore pull these coefficients well away from their priors (**Fig. S16**).

##### Relative reporting ratio (primary vs secondary infections)

The relative reporting ratio *γ_d_* quantifies the probability of reporting a primary infection relative to a secondary infection, with *γ_d_* < 1 expected in dengue-endemic settings where secondary infections present more severe clinical manifestations.

We model *γ_d_* using a hierarchical structure with province-level means and district-level deviations:

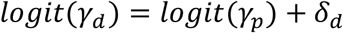

where *γ_p_* is the province-level mean on the probability scale and *δ_d_*∼*Normal*(0, *σ_γ_*_,*p*_) is a district-level deviation on the logit-scale. The priors are:

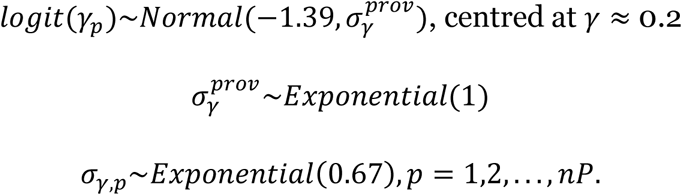

The prior mean of approximately 0.2 reflects the expectation that primary infections, which are typically milder, are reported at roughly one-fifth the rate of secondary infections. The province-specific scale parameter *σ_γ_*_,*p*_ allows the degree of within-province heterogeneity in *γ* to vary across provinces.

#### Seroprevalence survey data integration

In addition to case surveillance data, the model jointly fits age-stratified seroprevalence surveys conducted at any time point, including years before the surveillance data period. For a survey of individuals aged *A* years at the start of calendar year *Y*, cumulative FOI exposure is computed by summing annual FOI from birth year (*Y* − *A*) through year *Y* − 1:

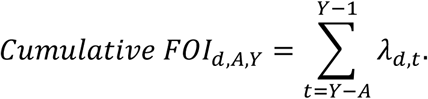

The survey year itself is excluded, consistent with the assumption that age *A* represents *A* complete years of exposure (i.e., exposure spans from the birth year through the year before the survey). For individuals with birth years prior to the surveillance data period (i.e., *Y* − *A* < first surveillance year), historical FOI estimates are used for the corresponding years. This allows surveys conducted prior to the surveillance data period, such as the 2014 nationwide urban serosurvey, to contribute to inference by constraining historical FOI trajectories.

Predicted seroprevalence for individuals aged *A* in survey *s* is then calculated as:

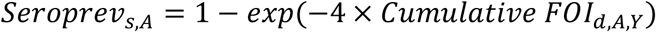

where the factor of 4 accounts for the assumed equal transmission risk to four co-circulating dengue serotypes. For age group *g* spanning ages *g*_1_ to *g*_2_ (with *n_g_* = *g*_2_ − *g*_1_ + 1), seroprevalence is averaged across individual ages:

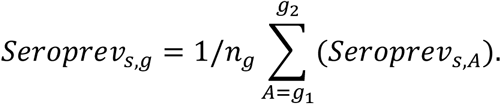

The likelihood for seroprevalence data follows a binomial distribution:

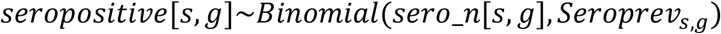

where *seropositive*[*s*, *g*] is the number of seropositive individuals from survey *s* in age group *g*, and *sero*_*n*[*s*, *g*] is the corresponding sample size.

#### Likelihood

The model accommodates three data types within a single likelihood, with separate contributions from age-stratified case data, aggregate case data, and seroprevalence surveys.

##### Age-stratified case data

When age-stratified case counts are available for district *d* in year *t*, the likelihood contribution is a negative binomial term for each age group *g*:

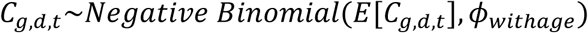

where *E*[*C_g_*_,*d*,*t*_] is the expected number of reported cases and *φ_wit_*_ℎ*age*_ is the overdispersion parameter.

##### Aggregate case data

When only aggregate case totals are available, the likelihood is a single negative binomial term for the total:

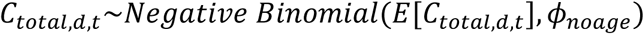

where *E*[*C_total_*_,*d*,*t*_] = ∑*_g_ E*[*C_g_*_,*d*,*t*_] and *φ_noage_* is a separate overdispersion parameter. Separate overdispersion parameters are used for the two data types because age-stratified counts, being split across age groups, are smaller in magnitude and may exhibit different variance structures compared to aggregate totals.

##### Seroprevalence data

For seroprevalence surveys, the likelihood follows a binomial distribution as described in the previous section:

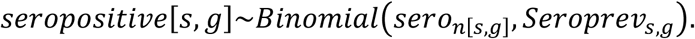

##### Missing data

District-year combinations with no reported data make no likelihood contribution. This design allows districts with limited or incomplete surveillance records to still benefit from shared information through hierarchical structures, national baseline, and covariate effects.

##### Overdispersion priors

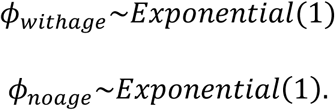

Weakly informative priors are placed on both overdispersion parameters.

#### Model scenarios

To evaluate the contribution of each information source to parameter estimation, we fitted four model scenarios to the data from Java and Bali, varying the inclusion of external covariates and seroprevalence data:

1. Full model: covariates (**Table 2**) and seroprevalence surveys included;
2. Seroprevalence only: seroprevalence surveys included, no covariates;
3. Covariates only: covariates included, no seroprevalence surveys; and
4. Case data only: neither covariates nor seroprevalence surveys included.

All four scenarios retain the same hierarchical structure, national baseline, AR(2) district residuals and reporting rate model. In scenarios without covariates, the covariate terms are removed from the FOI equation and the surveillance capacity terms are removed from the reporting rate equation, so that spatial heterogeneity is captured entirely by the hierarchical random effects. In scenarios without seroprevalence, the binomial likelihood component is omitted and inference relies solely on case surveillance data.

This comparison allows us to assess whether covariates and seroprevalence data improve identifiability of FOI and reporting parameters, particularly in districts with aggregate-only case data where FOI and reporting rate may be weakly identifiable from case counts alone.

#### Synthetic data validation

##### Data generation

To validate the model’s ability to recover known parameter values under varying data completeness, we conducted a simulation study. We generated synthetic data for 50 districts (5 provinces x 10 districts per province) over 9 years, mirroring the structure of the real data. The data generation procedure was as follows:

1. A national temporal baseline FOI trajectory (*β*_0,*t*_) was generated assuming multi-annual outbreak patterns.
2. For each district, covariate values were independently drawn from standard normal distributions (mean 0, SD 1) at both district and province levels, with fixed true coefficients used to generate heterogeneity in FOI and reporting rates.
3. District-specific FOI (*λ_d_*_,*t*_) was computed using the model’s generative model structure:

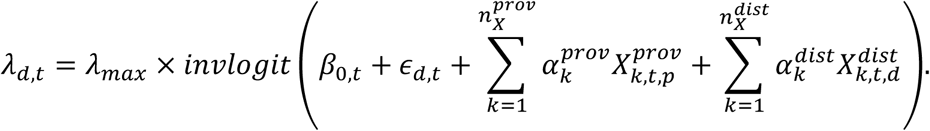
4. Reporting rates (*ρ_d_*) were generated from the reporting rate model:

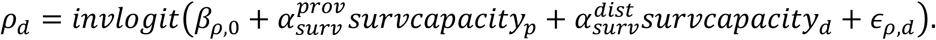
5. Relative reporting ratios (*γ_d_*) were simulated assuming an average of 0.2 across districts.
6. Population immunity and expected cases by single year of age were computed using the same annual update equations, historical FOI structure and population weighting as the model.
7. Age-stratified cases counts were simulated using negative binomial random draws for each district-year-age group combination.

The variance of the AR(2) residuals was set so that covariates explained approximately 10% of the between-district variation in FOI, comparable to the share estimated for the Java and Bali data. This setting was used for the main analysis, and settings in which covariates explained approximately 50% and 90% of this variation were used in the sensitivity analyses.

##### Data completeness scenario

We simulated four scenarios of decreasing data completeness to evaluate the model’s robustness when age-stratified surveillance data are limited:

1. S1: All 50 districts provide 9 age groups;
2. S2: All 50 districts provide 4 age groups;
3. S3: Provinces 1–2: 9 age groups; provinces 3–5: 4 age groups; and
4. S4: Province 1: 9 age groups; province 2: 4 age groups; provinces 3–5: aggregate only.

Scenario S4 most closely mirrors the heterogeneity in data availability observed in real data from Indonesia (**Table 1**). Each scenario was fitted both with and without covariates, yielding eight model runs in total.

##### Synthetic seroprevalence surveys

To assess the contribution of seroprevalence data, each scenario was also refitted with a simulated serosurvey in 10 districts (2 per province), covering ages 1-18 years with sample sizes per age matching the 2014 nationwide survey. The survey was placed either at the end of 2014, before the surveillance period, or at the end of 2019, within it.

##### Covariate misspecification

To assess robustness to misspecified covariates, each scenario was refitted with an additional district-level covariate drawn as an independent noise, unrelated to FOI, and with a partial covariate set in which on province-level and one district-level covariate used to generate the data were omitted.

##### Validation metrics

We compared posterior medians and 90% credible intervals of FOI (*λ_d_*_,*t*_), reporting rates (*ρ_d_*), and covariate coefficients (*α_k_*) to the true parameter values used in simulation, together with seroprevalence at ages 5, 9 and 15 at the end of 2024. We computed root mean squared error (RMSE) and 90% credible interval coverage for each parameter class. We also assessed how accuracy varies over the surveillance period by computing RMSE of seroprevalence at age 9 at the end of each year from 2016 to 2024.

#### Computational implementation

The model is implemented in the Stan probabilistic programming language and compiled via the cmdstanr package in R (*37*, *38*). Posterior inference is performed using Hamiltonian Monte Carlo with the No-U-Turn Sampler. We ran four chains, each with 2,000 warmup and 2,000 sampling iterations, using an adaptation target acceptance rate (*adapt*_*delta*) of 0.95 and maximum tree depth of 12, with a fixed random seed. In the real-data fits, R-hat values were at or below 1.01, bulk and tail effective sample sizes exceeded 400 for all quantities, and each model had at most three divergent transitions out of 8,000 draws. The energy Bayesian fraction of missing information (E-BFMI) was below the conventional threshold of 0.3 in all chains (0.23-0.30), associated with the overdispersion parameter for aggregate case data, but chains agreed closely (**Table S2**).

To improve sampling efficiency for hierarchical parameters, we employ non-centred parameterisations throughout. Large intermediate matrices (*S*(*a*, *d*, *t*), *M*(*a*, *d*, *t*), *I*_1_(*a*, *d*, *t*), and *I*_2_(*a*, *d*, *t*)) are declared as local variables within the model block and not stored in the output, reducing memory usage and output file size. These quantities can be reconstructed from the saved parameter posterior samples.

#### Post-hoc analyses

##### Model comparison

We compared the four model scenarios using approximate leave-one-out-cross-validation (LOO-CV) via Pareto-smoothed importance sampling (PSIS-LOO (*39*)). Expected log pointwise predictive density (ELPD) differences between models were computed from generated quantities in Stan. We assessed the reliability of LOO estimates using Pareto-k diagnostics, with k > 0.7 indicating potentially unreliable estimates for individual observations. LOO-CV was performed separately for case-only and joint case-seroprevalence likelihoods to evaluate model fit under both data configurations.

##### Seroprevalence prediction and vaccine eligibility

Using posterior FOI samples from the full model, we predicted age-specific seroprevalence for each district at the end of 2024 by computing cumulative exposure from birth year through 2024. For children aged 9 years, this exposure (2016-2024) lies entirely within the surveillance period.

The WHO 2024 position paper on dengue vaccines recommends the use of TAK-003 (Qdenga) in children aged 6–16 years in settings with high dengue transmission intensity (*5*). To determine transmission intensity, the WHO suggests that a dengue seroprevalence exceeding 60% by age 9 could be considered an indicator of high transmission, while noting that threshold cut-offs should be decided by individual countries. We adopted this criterion to assess vaccine eligibility across districts. For each district, we estimated the median posterior seroprevalence at age 9 and computed the posterior probability that this seroprevalence exceeds 60%, *P*(*seroprevalence_d_*_,*A*=9_ > 0.60). Districts were classified as high-priority candidates for vaccine introduction when this posterior probability exceeded 75%, providing a more conservative criterion that accounts for estimation uncertainty.

##### Comparison with incidence-based prioritisation

To assess whether routine surveillance data alone could identify the same high-priority districts, we compared the model-based seroprevalence classification against a simpler incidence-based criterion, which selected the *K* districts with the highest mean reported incidence rate over 2016-2024, where *K* is the number of districts classified as high priority by the model. We evaluated agreement using the strict criterion (median seroprevalence at age 9 > 60% and posterior probability > 75%) and the lenient criterion (median > 60%), and compared the number of districts selected by both approaches with that expected if *K* districts were selected at random.

## Data Availability

The routine surveillance data for Jakarta are freely accessible from: https://surveilans-dinkes.jakarta.go.id/. Researchers interested in accessing additional surveillance data may contact the Arbovirus control unit of the Vector-Borne and Zoonotic Diseases, Venomous Animal Bites, and Poisonous Plants working team of the Ministry of Health at

## Authors’ contributions

B.A.D.: conceptualization, funding acquisition, formal analysis, methodology, writing— original draft; I.R.F.E.: writing—review and editing; A.S.: writing—review and editing; F.S.M.S.: data curation, writing—review and editing; A.H.: data curation, writing—review and editing; B.T.: writing—review and editing; D.A.: data curation, writing—review and editing; D.G.: writing—review and editing; H.H.: writing—review and editing; A.K.N.: writing—review and editing; E.P.: writing—review and editing; I.S.: writing—review and editing; A.R.C.: writing—review and editing; A.T.H.: writing—review and editing; H.C.: writing—review and editing; S.B.: writing—review and editing; S.M.: conceptualization, funding acquisition, methodology, writing—review and editing. All authors gave final approval for publication.

## Conflict of interest declaration

We declare we have no competing interests.

### Acknowledgements

We thank the Vector-Borne and Zoonotic Diseases, Venomous Animal Bites, and Poisonous Plants Working Team at the Ministry of Health of Indonesia for providing access to the surveillance data and their technical support.

## Data and code availability

The codes to perform analyses in this article available at: https://github.com/mlgh-sg/dengue-covariates-catalytic. The routine surveillance data for Jakarta are freely accessible from: https://surveilans-dinkes.jakarta.go.id/. Researchers interested in accessing additional surveillance data may contact the Arbovirus control unit of the Vector-Borne and Zoonotic Diseases, Venomous Animal Bites, and Poisonous Plants working team of the Ministry of Health at

