## Supplementary material for "Bridging surveillance gaps in dengue: a hierarchical model integrating mixed data sources for transmission estimation and vaccine targeting": SI

### SUPPLEMENTARY MATERIALS

This is the supplementary materials for the manuscript: "***Bridging surveillance gaps in dengue: a hierarchical model integrating mixed data sources for transmission estimation and vaccine targeting***".

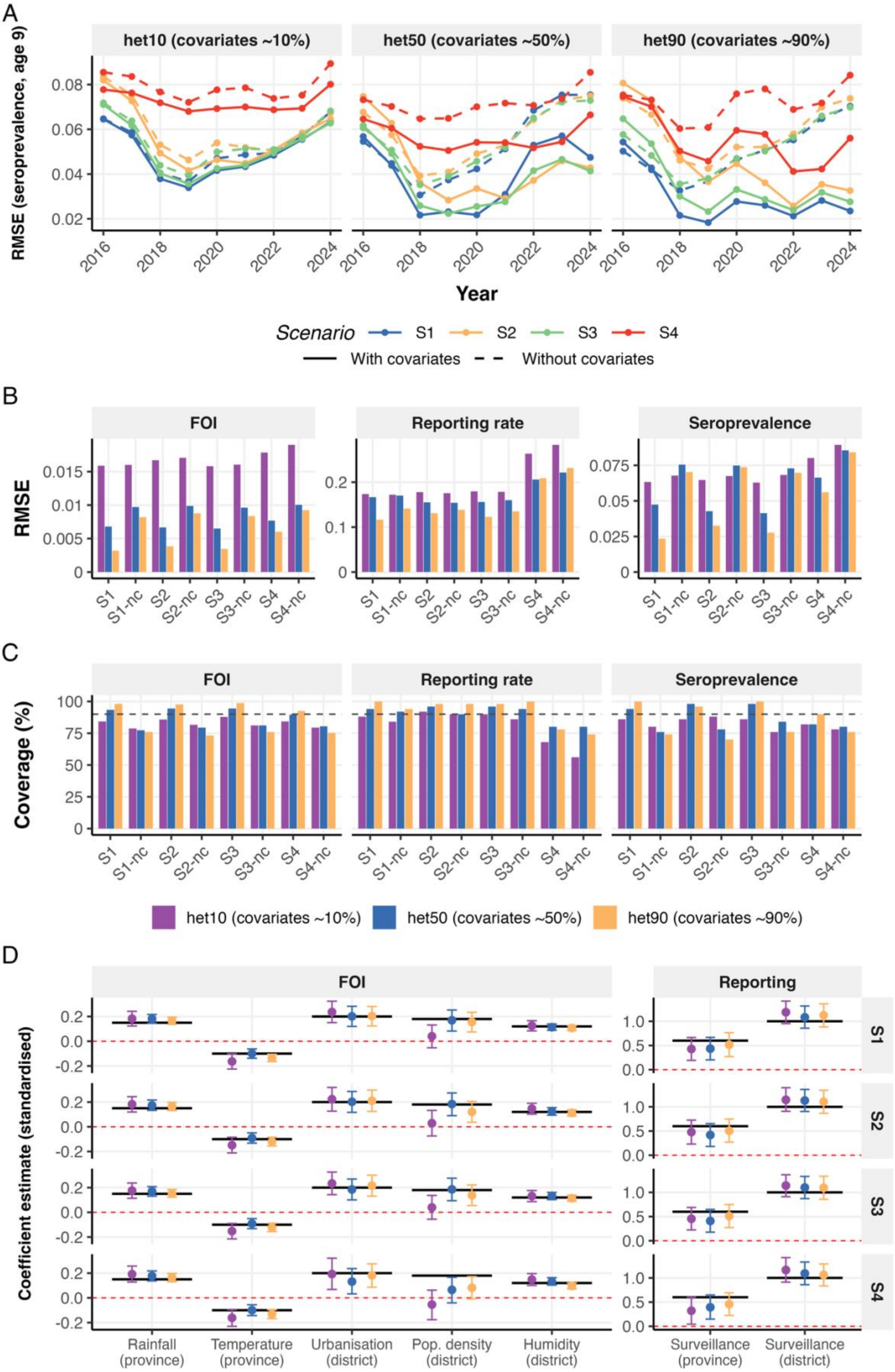
A
het10 (covariates ~10%)
het50 (covariates ~50%)
het90 (covariates ~90%)
RMSE (seroprevalence, age 9)
0.02
0.04
0.06
0.08
2016
2018
2020
2022
2024
Year
Scenario
S1
S2
S3
S4
With covariates
Without covariates
B
FOI
Reporting rate
Seroprevalence
RMSE
0
0.005
0.01
0.015
0.1
0.2
0.025
0.05
0.075
S1
S1-nc
S2
S2-nc
S3
S3-nc
S4
S4-nc
C
Coverage (%)
25
50
75
100
het10 (covariates ~10%)
het50 (covariates ~50%)
het90 (covariates ~90%)
D
FOI
Reporting
Coefficient estimate (standardised)
-0.2
0.0
0.2
0.5
1.0
Rainfall (province)
Temperature (province)
Urbanisation (district)
Pop. density (district)
Humidity (district)
Surveillance (province)
Surveillance (district)

**Figure S1. Synthetic validation across levels of covariate-explained FOI heterogeneity.** Covariates explain about 10% (het10, main design), 50% (het50) or 90% (het90) of the between-district variance in FOI. **(A)** RMSE of seroprevalence at age 9 at the end of each surveillance year, by scenario, with (solid) and without (dashed) covariates. **(B)** RMSE and **(C)** 90% credible interval coverage for FOI, reporting rate and seroprevalence at age 9, by scenario and heterogeneity level. **(D)** Recovery of FOI and reporting covariate coefficients (posterior median and 90% CrI). Black lines denote true simulated values, and red dashed lines mark zero. Dashed grey lines in **(C)** indicate the nominal 90% coverage level.

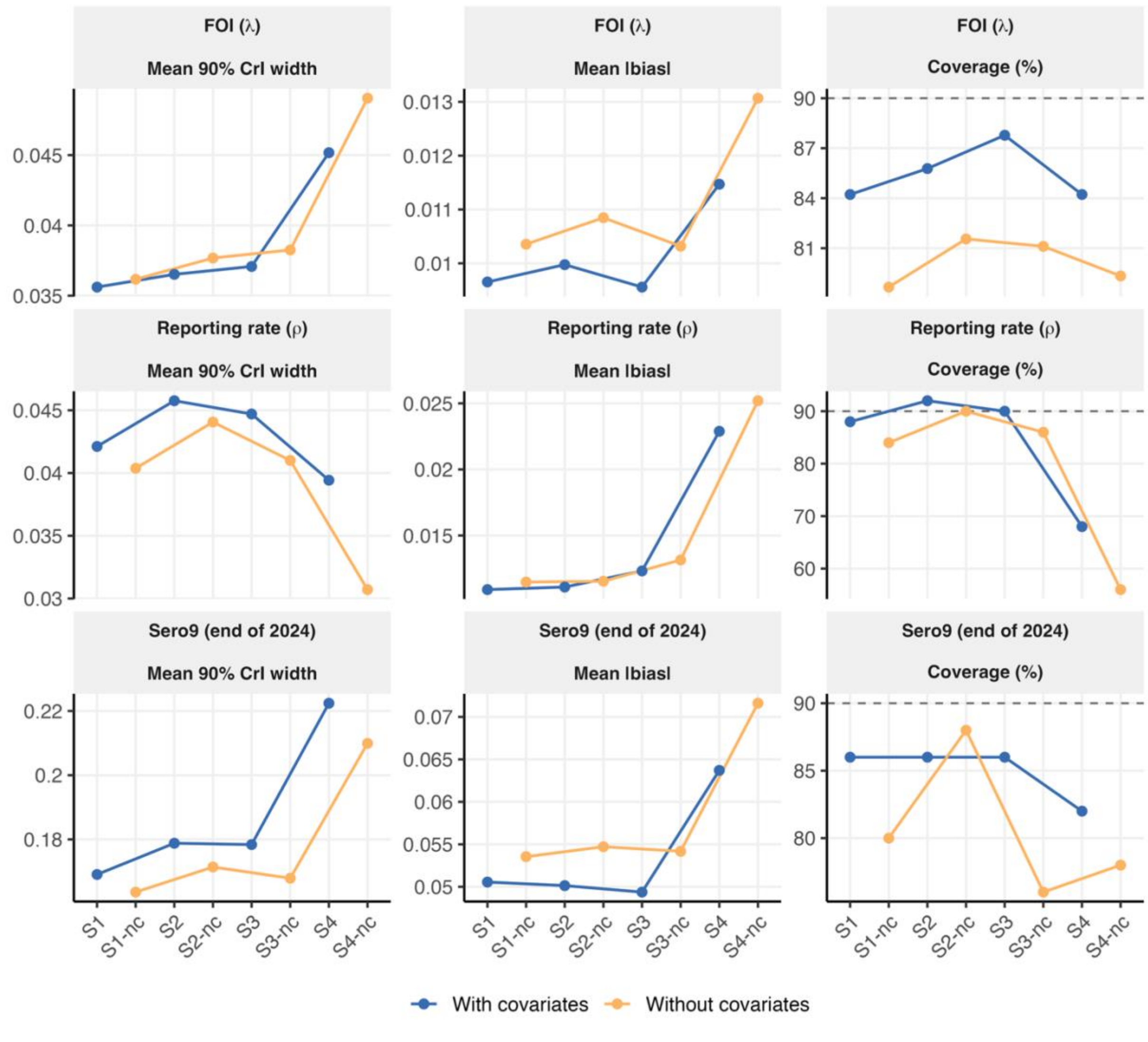


**Figure S2. Decomposition of estimation performance across data completeness scenarios without seroprevalence data (het10).** Mean 90% credible interval width (left), mean absolute bias (centre) and 90% credible interval coverage (right) for FOI, reporting rate, and seroprevalence at age 9 at the end of

2024, by scenario, with and without covariates (-nc). Dashed lines indicate the nominal 90% coverage level. In scenario S4, reporting rate intervals narrow while absolute bias increases, so the shortfall in coverage reflects overconfident shrinkage rather than wide posteriors.

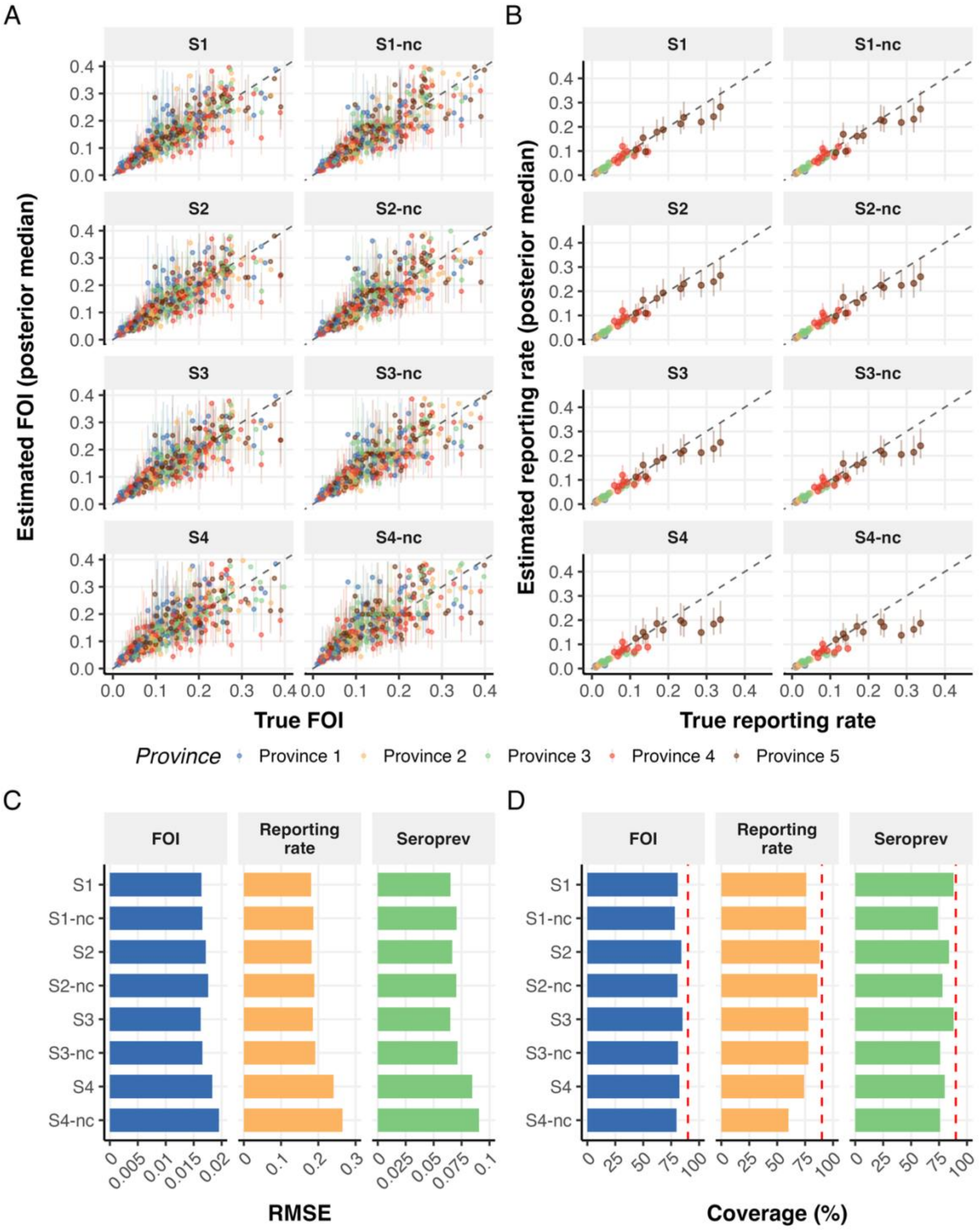

**Figure S3. Synthetic validation with a seroprevalence survey at the end of 2014 (before surveillance, matching the real data design). (A)** Estimated versus true FOI and **(B)** estimated versus true reporting rate by scenario and province, with 90% CrI. **(C)** RMSE and **(D)** 90% credible interval coverage for FOI, reporting rate and seroprevalence at age 9. Dashed red lines indicates the nominal 90% coverage level.

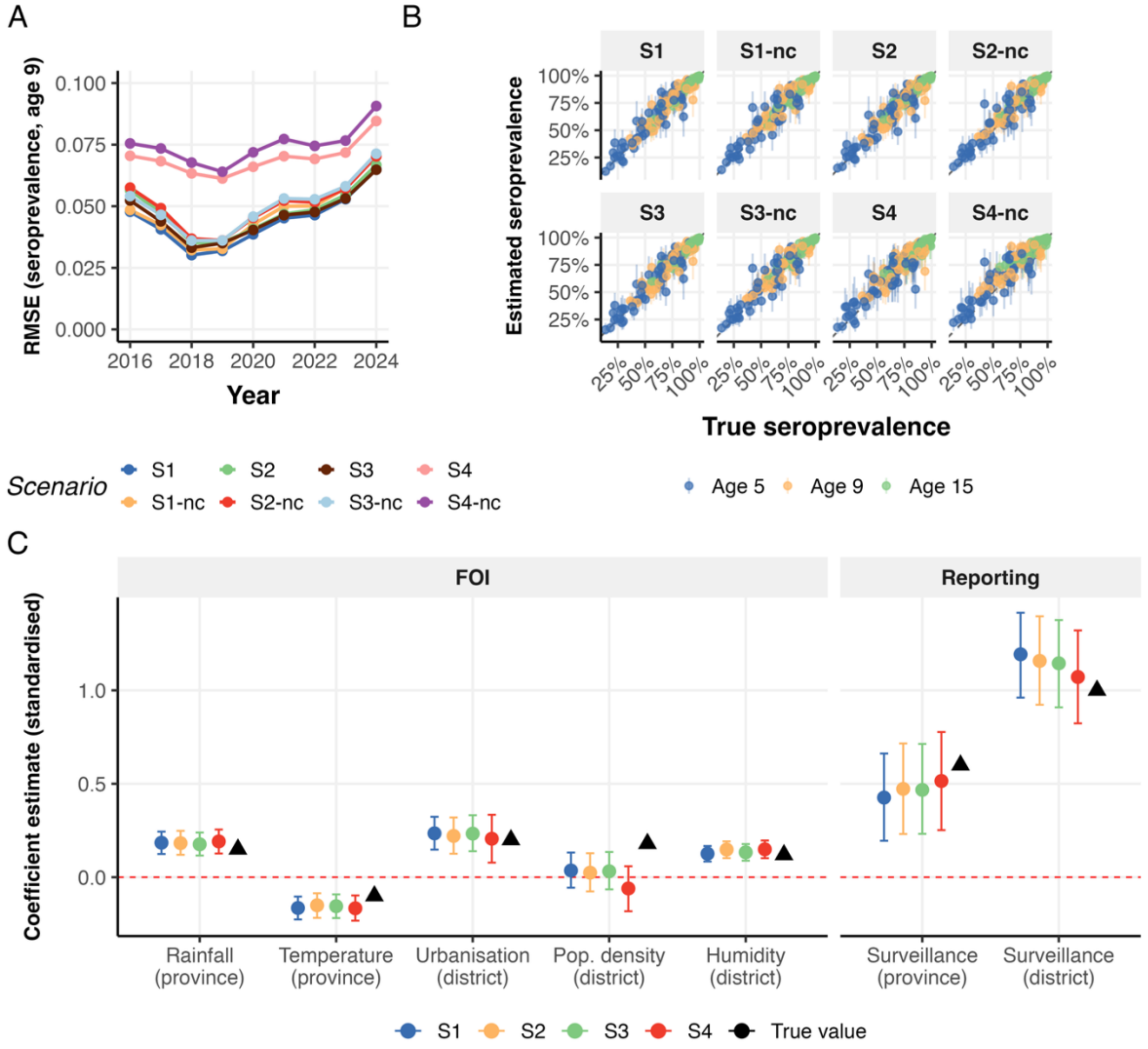


**Figure S4. Synthetic validation with a seroprevalence survey at the of 2014, showing seroprevalence and covariate recovery. (A)** RMSE of seroprevalence at age 9 at the end of each surveillance year. **(B)** Estimated versus true seroprevalence at ages 5, 9 and 15 at the end of 2024, by scenario. **(C)** Recovery of FOI and reporting covariate coefficients (posterior median and 90% CrI). Black triangles denote true simulated values.

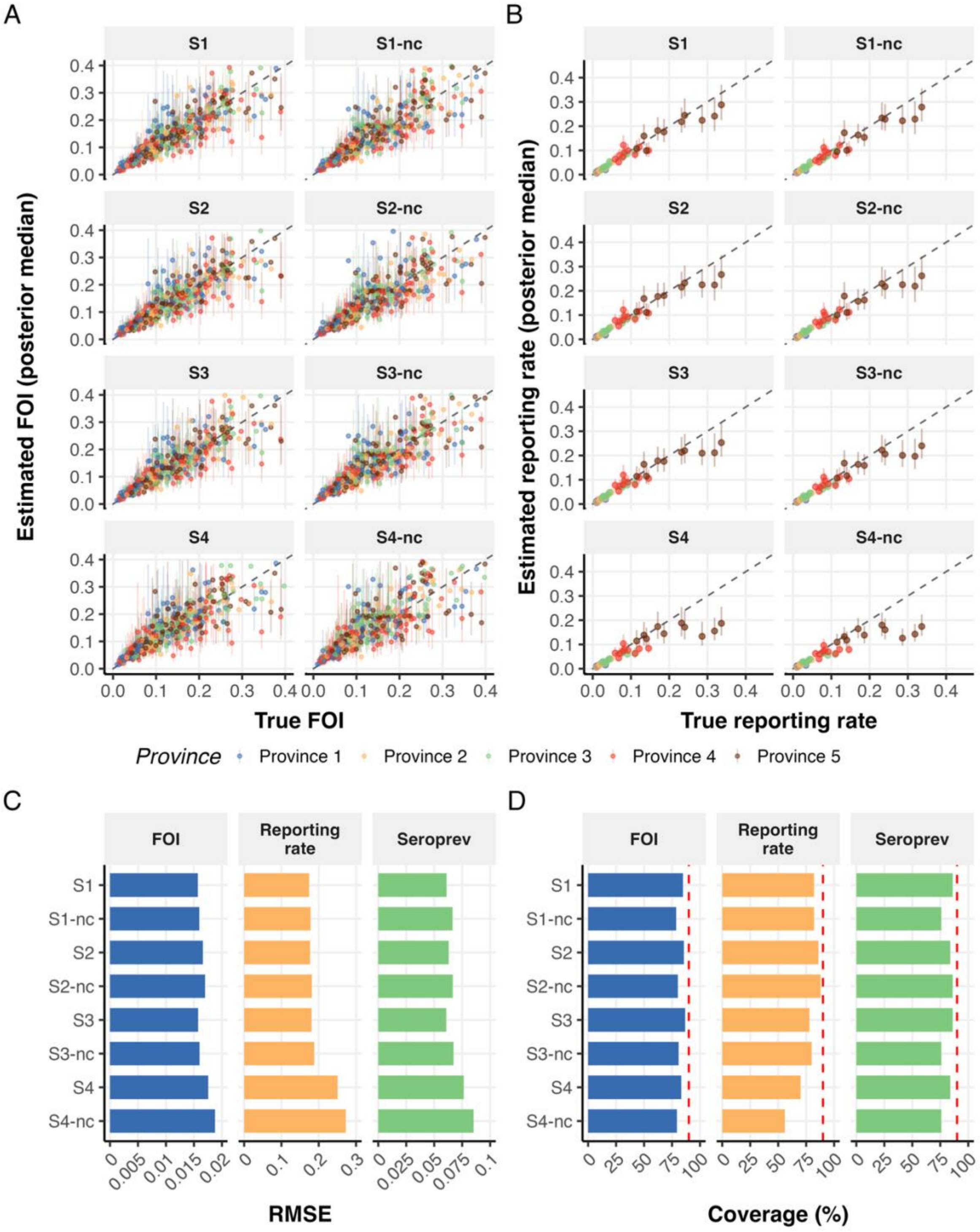


**Figure S5. Synthetic validation with a seroprevalence survey at the end of 2019 (mid-surveillance).** Panels as in **Fig. S3**.

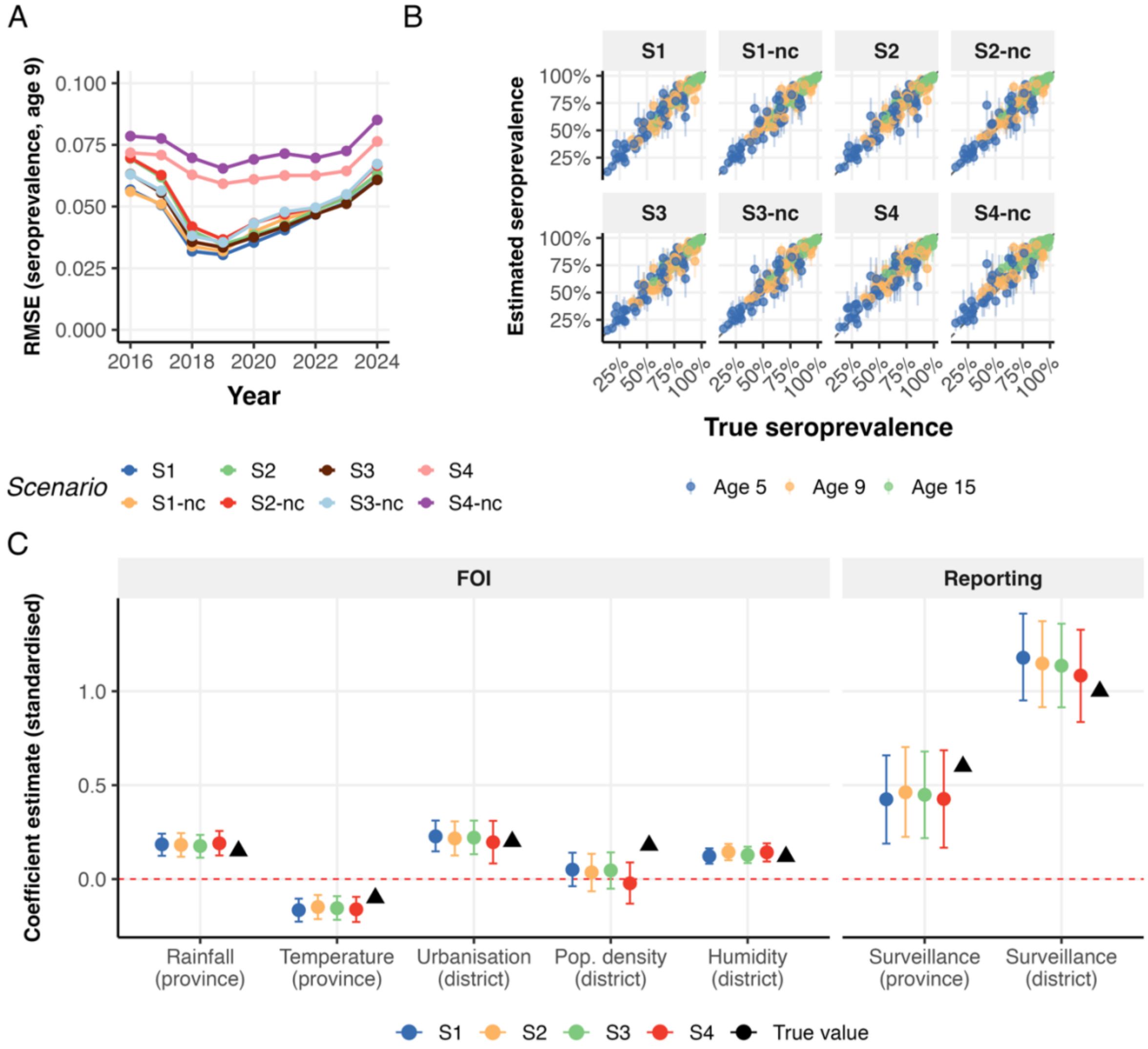


**Figure S6. Synthetic validation with a seroprevalence survey at the end of 2019, showing seroprevalence and covariate recovery.** Panels as in **Fig. S4.**

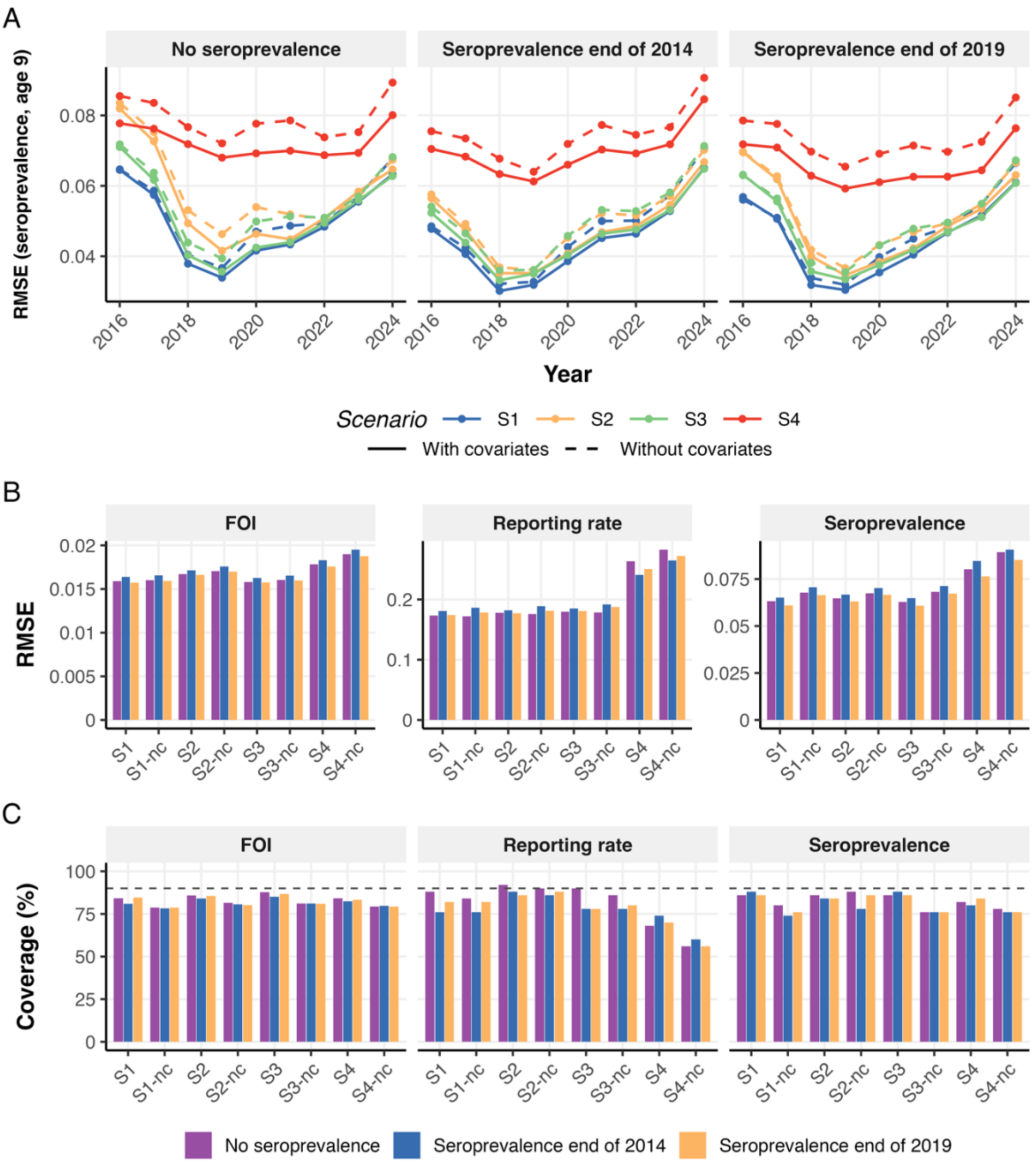


**Figure S7. Direct comparison of synthetic model performance without a seroprevalence survey, with a serosurvey at the end of 2014, and with a serosurvey at the end of 2019. (A)** RMSE of seroprevalence at age 9 over time by scenario, with (solid) and without (dashed) covariates. **(B)** RMSE and **(C)** 90% credible interval coverage for FOI, reporting rate and seroprevalence, by scenario and survey design. Dashed lines indicate the nominal 90% coverage level. A survey mainly improves seroprevalence estimates in the early surveillance years and changes little at the end of 2024, whereas reporting rate coverage decreases slightly when a survey is added.

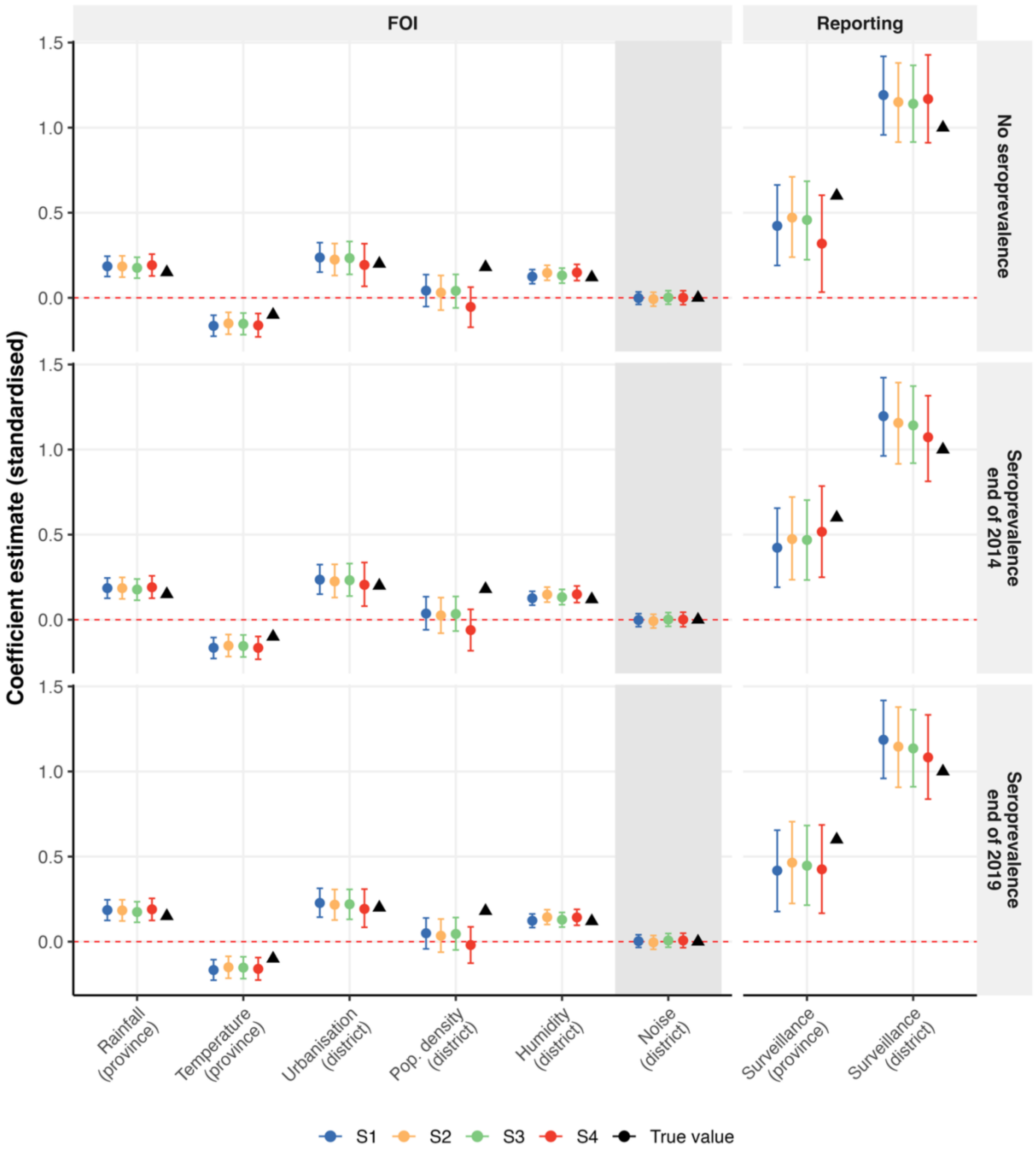


**Figure S8. Model robustness to inclusion of an uninformative covariate.** Posterior estimates (median and 90% CrI) of covariate coefficients when a synthetic noise covariate is added to the district-level FOI predictors, across all four synthetic data scenarios. The noise covariate is drawn from an independent standard normal distribution, with a true value of 0. Rows show fits without seroprevalence data and with a survey at the end of 2014 or 2019. Black triangles denote true values. The noise covariate coefficient (shaded column) remains centred near zero in all scenarios, and estimates for the genuine covariates are unaffected.

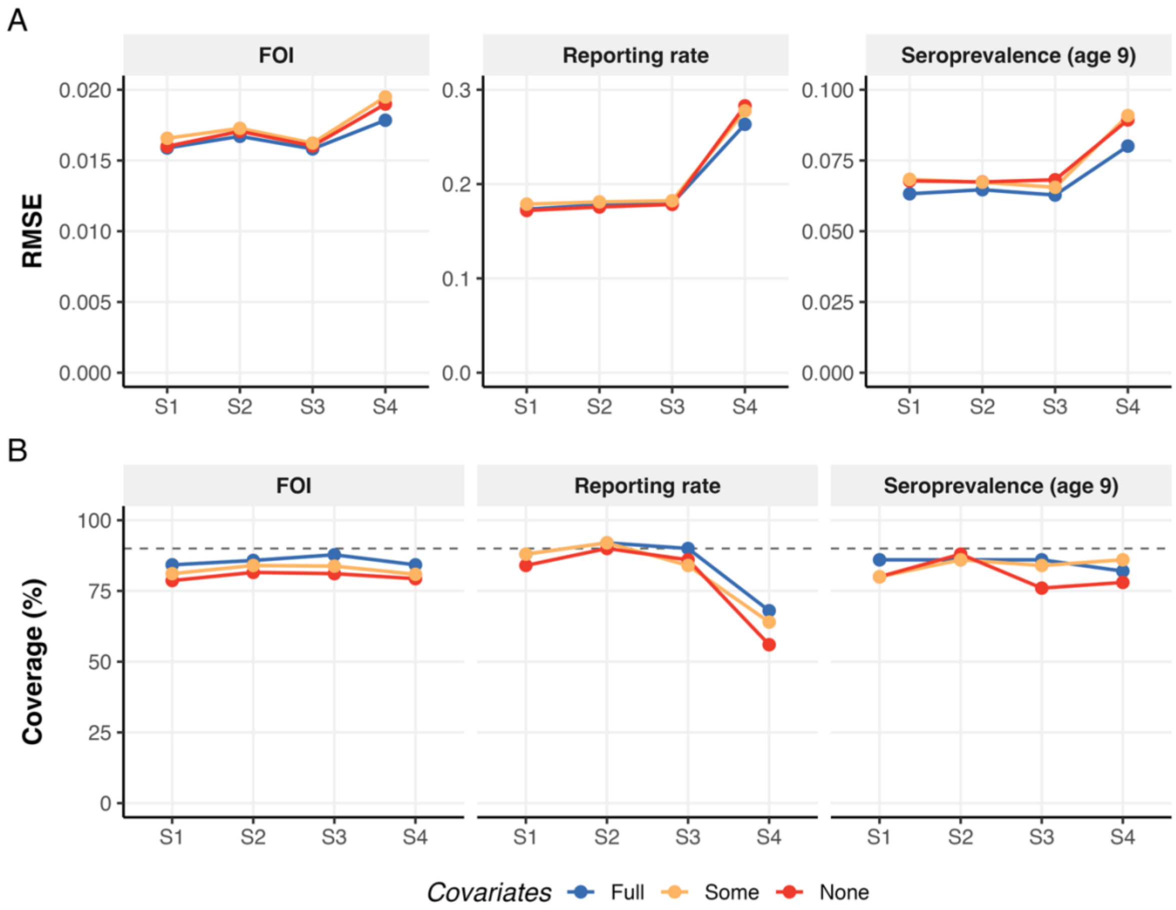


**Figure S9. Estimation performance with all, some or no covariates. (A)** RMSE and **(B)** 90% credible interval coverage for FOI, reporting rate and seroprevalence at age 9, compared with the true parameter values, for each data scenario. Dashed lines indicate the nominal 90% coverage level.

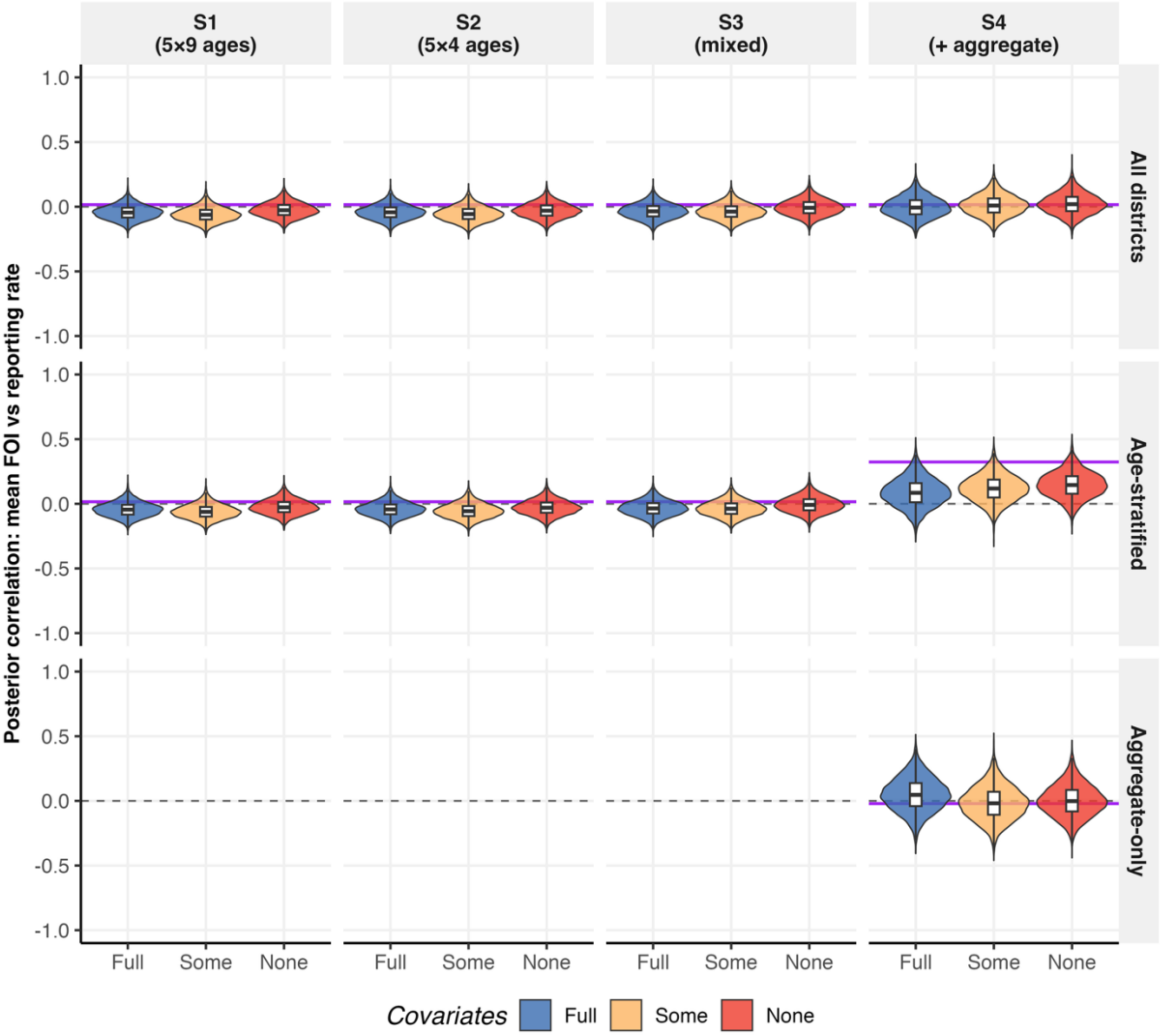


**Figure S10. Posterior correlation between district mean FOI and reporting rate for models with full, some and no covariates.** Violins show the posterior distribution of the correlation for each data scenario. Rows show all districts, districts with age-stratified data, and districts with aggregate data only (S4 only). Purple lines denote the correlation between the true parameters, and dashed lines mark zero.

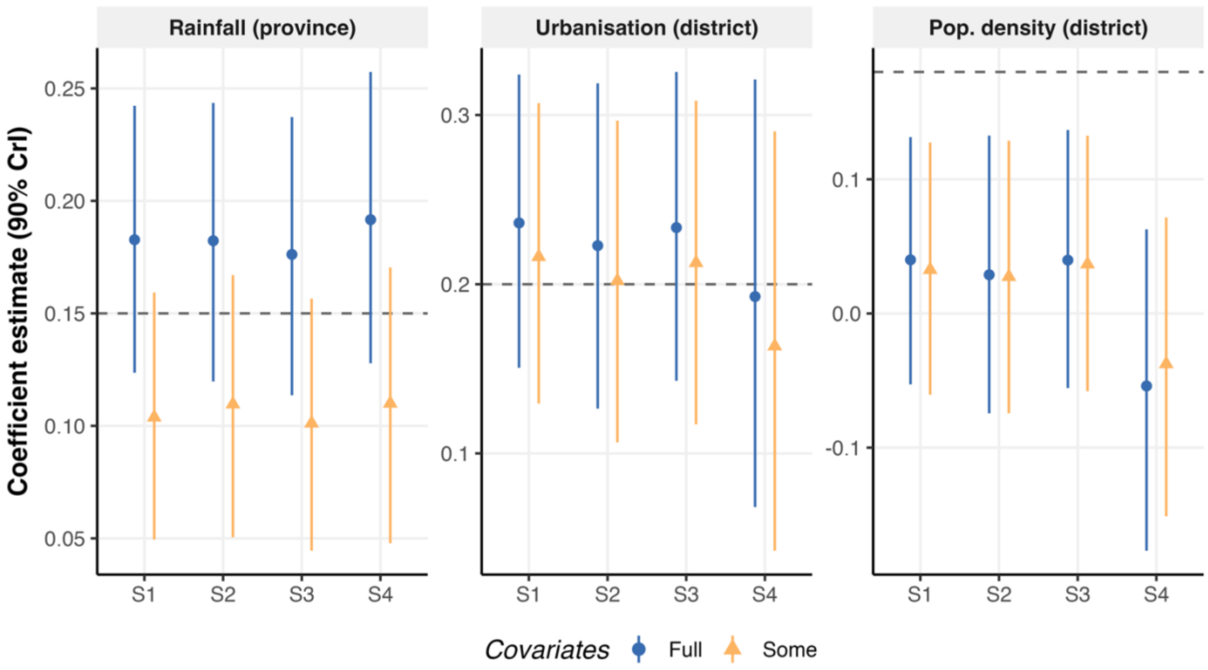


**Figure S11. Covariate coefficient estimates (posterior median and 90% CrI) for the covariates retained in the partial-covariate model.** Results from models with full and some covariates, for each data scenario. Dashed lines denote true simulated values.

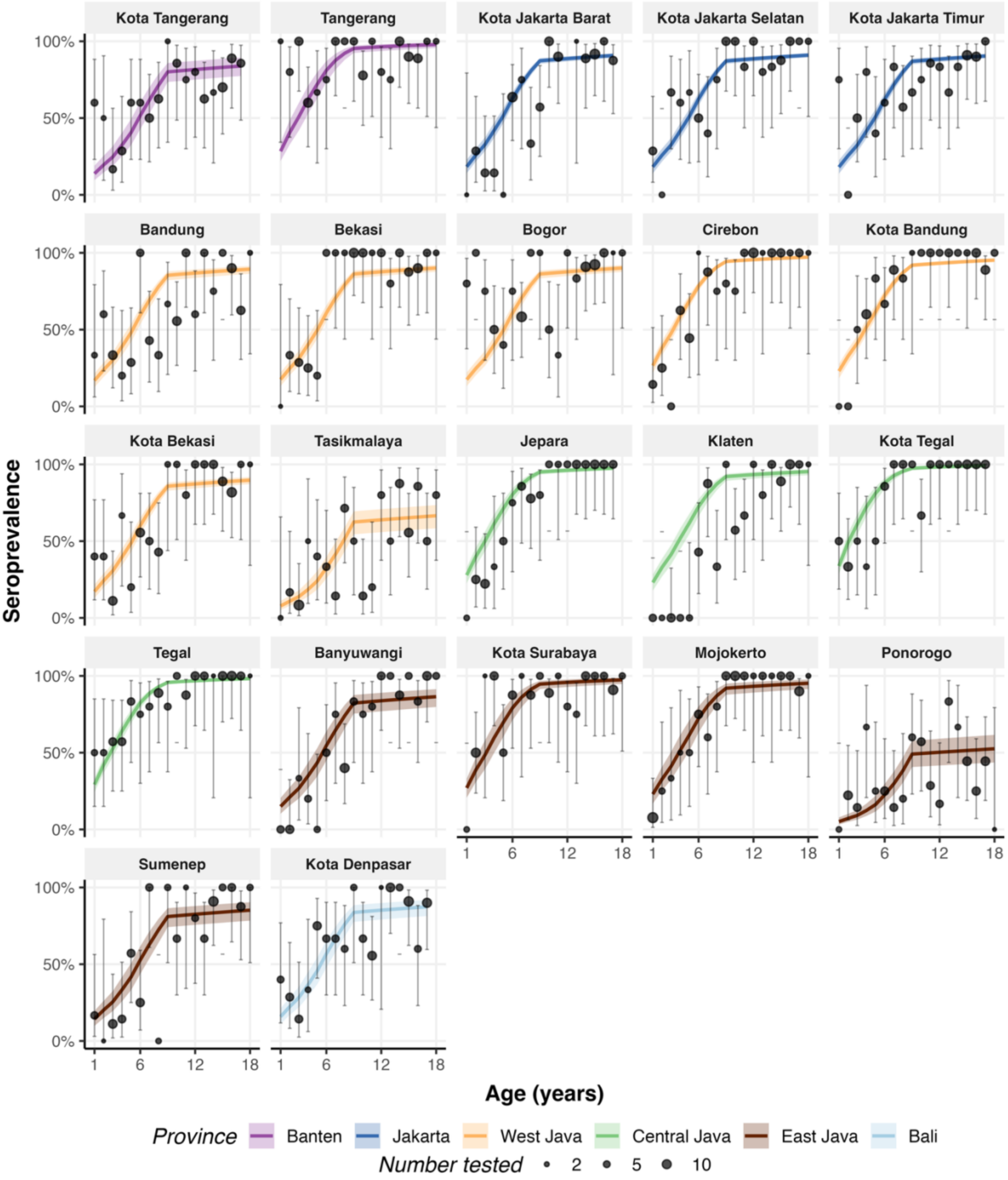


**Figure S12. Observed and fitted age-seroprevalence curves at the 22 sites of the 2014 national serosurvey.** Points show observed seroprevalence by single year of age with 95% Wilson confidence intervals, and point size is proportional to the number tested. Lines and ribbons show the full model posterior median and 90% CrI, coloured by province.

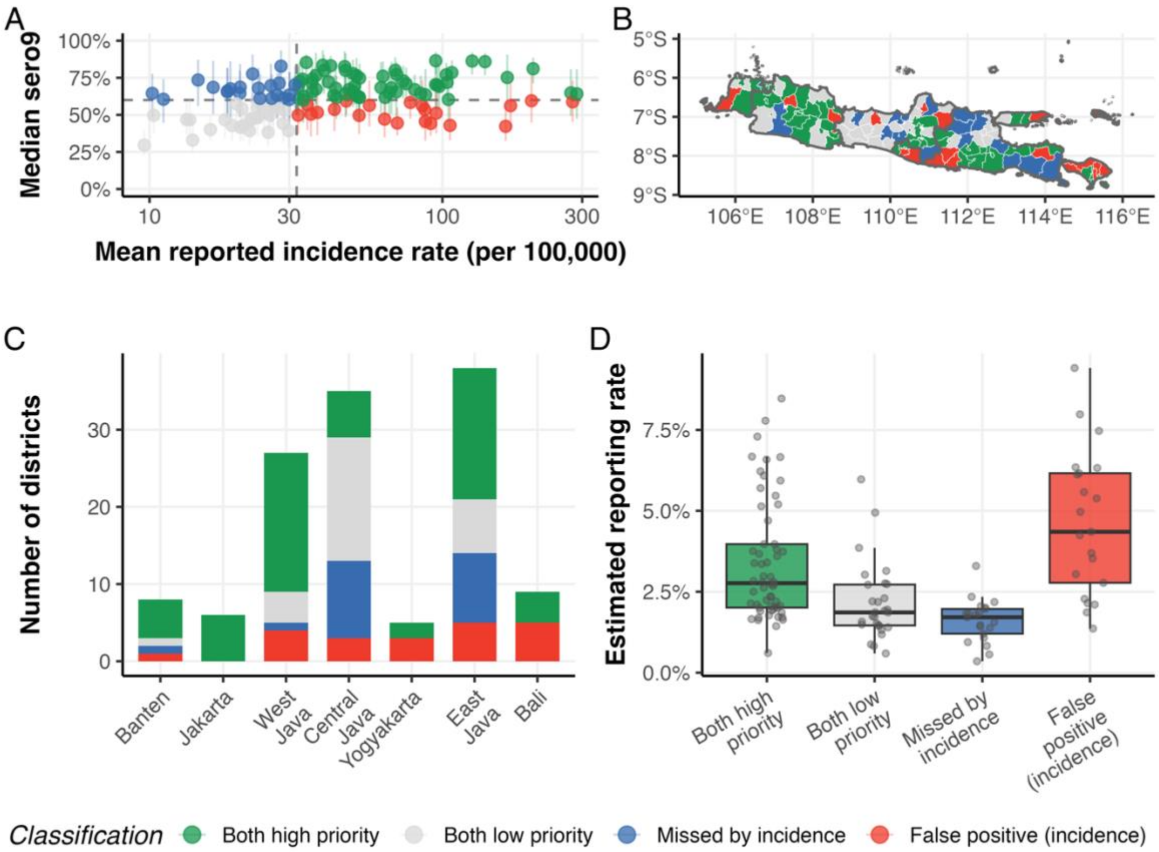


**Figure S13. Misalignment between model-based and incidence-based vaccine prioritisation with lenient criteria.** Model-based priority districts have median seroprevalence at age 9 above 60% at the end of 2024. Incidence-based priority districts are the same number of districts ranked highest by mean reported incidence. **(A)** Median seroprevalence at age 9 (with 90% CrI) versus mean reported incidence rate (per 100,000, log scale) by district. Dashed lines indicate the 60% seroprevalence threshold and the matched incidence threshold. **(B)** Spatial distribution of agreement categories across Java and Bali. **(C)** Number of districts in each agreement category by province. **(D)** Estimated reporting rate by agreement category.

96

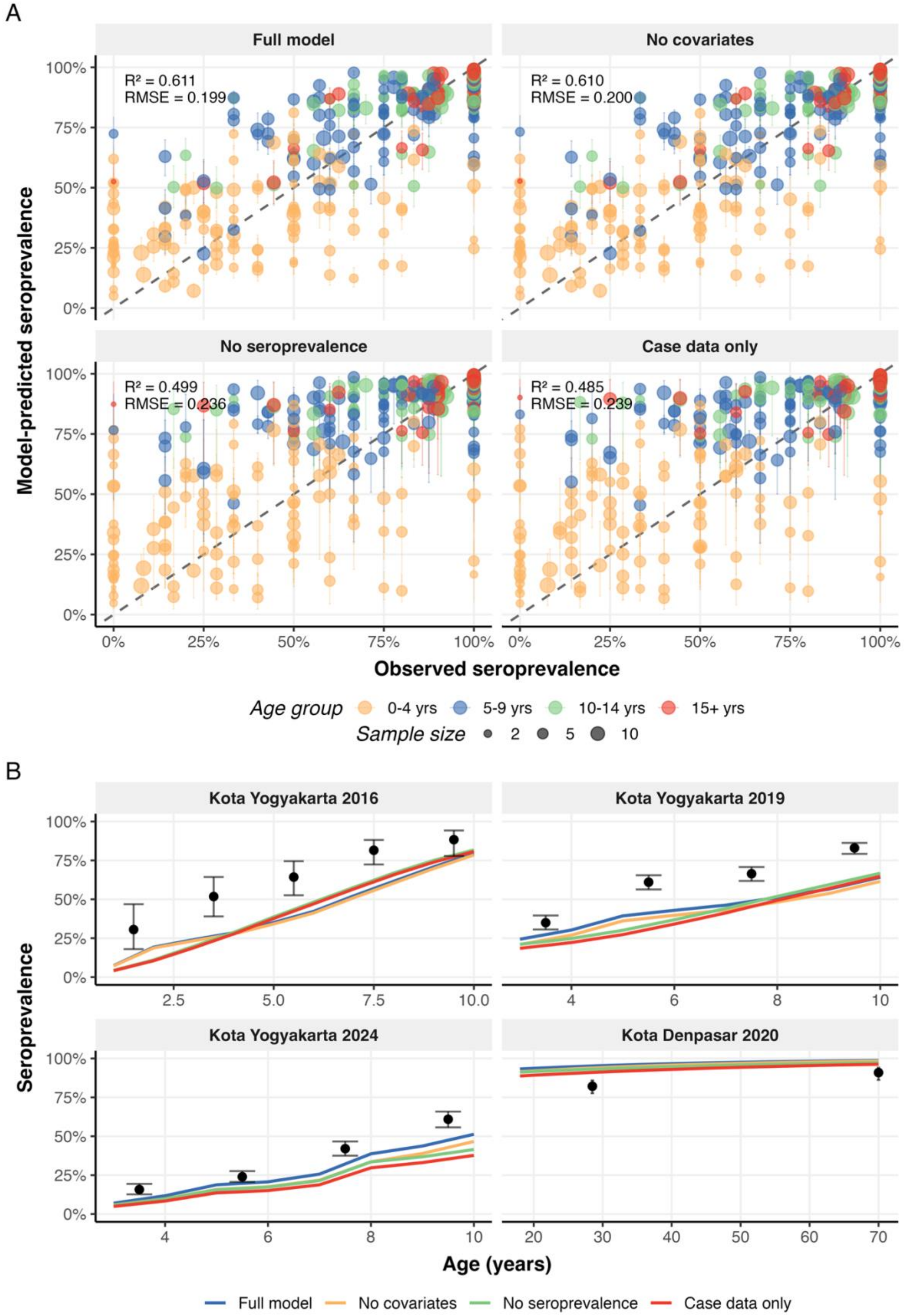
A
Full model
No covariates
No seroprevalence
Case data only
R² = 0.611
RMSE = 0.199
R² = 0.610
RMSE = 0.200
R² = 0.499
RMSE = 0.236
R² = 0.485
RMSE = 0.239
Model-predicted seroprevalence
Observed seroprevalence
Age group 0-4 yrs 5-9 yrs 10-14 yrs 15+ yrs
Sample size 2 5 10
B
Kota Yogyakarta 2016
Kota Yogyakarta 2019
Kota Yogyakarta 2024
Kota Denpasar 2020
Seroprevalence
Age (years)
Full model No covariates No seroprevalence Case data only

**Figure S14. Seroprevalence calibration across the four model variants. (A)** Model-predicted versus observed seroprevalence for the fitted 2014 national serosurvey, by age group, with $R^2$ and RMSE for each model. Point size reflects sample size, and error bars show 90% CrI. **(B)** Observed seroprevalence in the held-out surveys (Kota Yogyakarta 2016, 2019 and 2024; Kota Denpasar 2020) with 95% Wilson confidence intervals, and posterior median predictions from each model.

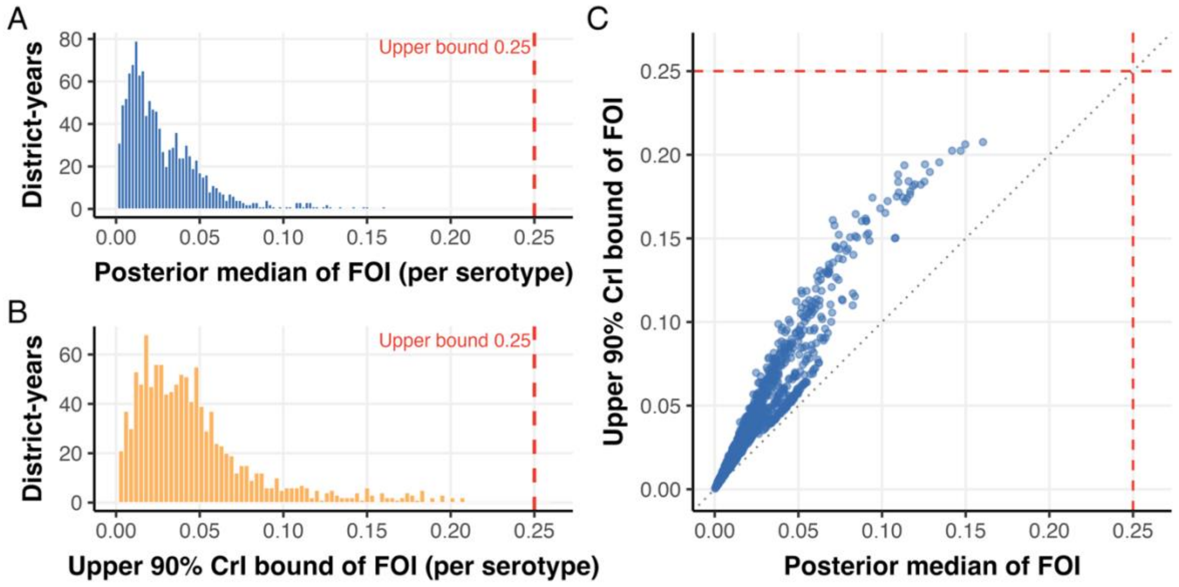


**Figure S15. Posterior estimates of district-year FOI relative to the upper bound $\lambda_{max}$.** Distribution of posterior medians (A), of upper 90% credible bounds (B), and their joint distribution (C). The dashed red line marks $\lambda_{max}$ = 0.25. The bound is not materially restrictive, as the largest upper 90% credible bound reaches 83% of $\lambda_{max}$.

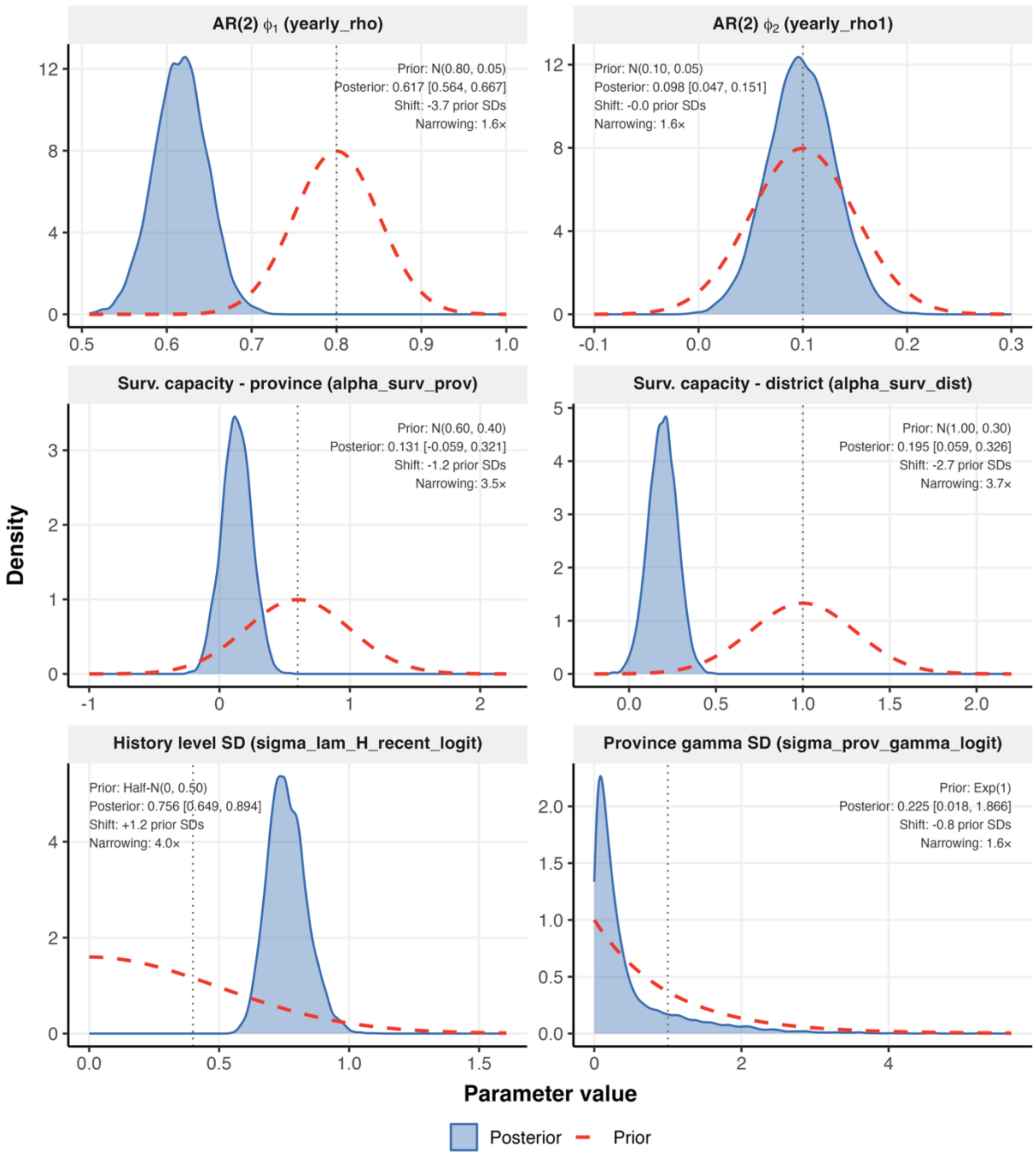


**Figure S16. Prior versus posterior distributions for selected parameters.** The panels show the AR(2) temporal coefficients, the province- and district-level surveillance capacity coefficients, the SD of the district historical FOI shift, and the SD of the province-level relative primary reporting effect. Dashed red curves show priors, shaded blue curves show posteriors, and dotted vertical lines mark the prior means. Annotations give each posterior median and 90% CrI, its shift in prior standard deviations and the narrowing relative to the prior. The first-order autoregressive coefficient, the district surveillance capacity coefficient and the historical FOI SD are informed by the data. The second-order autoregressive coefficient and the province-level reporting SD remain close to their priors.

**Table S1. Leave-one-out cross-validation comparison of the case data across four model scenarios.** Expected log predictive density (ELPD), its standard error, effective number of parameters (p_loo), LOOIC, and the number and percentage of observations with Pareto k > 0.7. ELPD differences and their standard errors are relative to the best model (case data only). The last three columns repeat the comparison using only observations with Pareto k ≤ 0.7 in all models.

| Model | ELPD (SE) | ΔELPD (SE) | ΔELPD, Pareto k ≤ 0.7 (SE) | Pareto k > 0.7 (%) |
|---|---|---|---|---|
| Case data only | -12,908.9 (86.1) | 0 | 0 | 21.5 |
| No seroprevalence | -12,912.5 (86.1) | -3.6 (5.4) | -1.0 (3.1) | 20.6 |
| No covariates | -12,950.8 (86.2) | -41.9 (6.2) | -23.5 (3.7) | 20 |
| Full model | -12,955.8 (86.1) | -46.9 (7.3) | -26.1 (5.2) | 19.4 |

**Table S2. MCMC sampler diagnostics for the four real-data models.** Maximum $\hat{R}$ and the number of parameters with $\hat{R}$ > 1.01, minimum bulk and tail effective sample size, number of divergent transitions and of iterations reaching the maximum tree depth, and the range of E-BFMI across the four chains, with the number of chains below 0.3. Each model used 4 chains with 2,000 warm-up and 2,000 sampling iterations.

| Model | Max $\hat{R}$ | Min bulk ESS | Min tail ESS | Divergent transitions | E-BFMI (range across chains) |
|---|---|---|---|---|---|
| Full model | 1.004 | 473 | 968 | 1 | 0.28-0.30 |
| No covariates | 1.011 | 435 | 680 | 3 | 0.25-0.27 |
| No seroprevalence | 1.012 | 470 | 928 | 0 | 0.24-0.27 |

| | | | | | |
|---|---|---|---|---|---|
| Case data only | 1.009 | 412 | 769 | 1 | 0.23-0.26 |
